# Identifying therapeutic targets for rheumatoid arthritis by genomics-driven integrative approaches

**DOI:** 10.1101/2024.03.19.24304536

**Authors:** Jie Zhang, Xinyu Fang, Jingwei Wu, Zixing Zhang, Min Mu, Dongqing Ye

## Abstract

Genomics-driven drug discovery framework holds promise in developing novel therapeutic targets. Here, we leveraged large-scale genomic data including genome-wide association studies (GWAS), rare variant burden tests in exome sequencing studies (Exome), and protein quantitative trait loci (pQTL), to prioritize potential therapeutic targets and identify opportunities for drug repositioning in rheumatoid arthritis (RA). We found that prioritized genes covering two approved RA treatment targets (IL6R and CD86), and five targets tested in clinical trials for RA. Eighteen proteins were identified as having causalities with RA risk, three out of them showed strong support for colocalization. Bromodomain-containing protein 2 (BRD2) was nominated as one of the most promising candidates for clinical translation as its wide expression in joint synovial tissues and validation in observational analyses associating with RA incidence. Collectively, our systematic screening of candidate drug targets from different genetically informed approaches, and provided a comprehensive insight into therapeutic strategies for RA.

## Introduction

Rheumatoid arthritis (RA), a common systemic autoimmune disease primarily characterized by chronic tissue inflammation and joint destruction,^1,2^ has an estimated heritability of nearly 50%.^3^ In 2020, an estimated 17.6 million people had RA worldwide.^4^ Over recent decades, advancements in the treatment of RA have been notable, with the advent of biologic disease-modifying anti-rheumatic drugs (bDMARDs) and, more recently, targeted synthetic DMARDs.^5^ Nevertheless, despite the availability of these therapeutic drugs for RA, a considerable portion of RA patients fail to achieve low disease activity or remission.^6^ Additionally, there exists a substantial cost burden and disparities in these medications accessibility,^7,8^ further complicating the situation. Currently, screening for novel therapeutic targets remains a pressing need in the management of RA.

Using human genetic data to optimize the selection of targets has potential to lead to approved drugs.^9,10,11^ A previous genome-wide association study (GWAS) meta-analysis for RA with >100,000 subjects,^3^ served as a paradigm, underscoring the pivotal role of genetics in drug discovery efforts, which revealed drug target genes of approved RA drugs significantly overlapped with the 98 identified biological candidate genes, and indicated that *CDK6*/*CDK4* inhibitors^12^ (approved drugs for different types of cancer) may hold promise for drug repositioning in RA treatment. One constraint inherent in the singular approach of drug discovery driven by GWAS lies in the fact that a significant portion of GWAS signals stemming from common variants manifest negligible individual effect magnitudes. However, their potential influence on molecular phenotypes, encompassing gene expression and protein abundance, might be significant to justify consideration for therapeutic intervention.^13^ Coupling large-scale GWAS with molecular quantitative trait loci (mQTL) datasets by utilizing Mendelian randomization studies, transcriptome-wide association studies, and colocalization methods provides insight to pathogenesis and facilitates future drug target prioritization.^14,15,16,18,19^ Besides, GWAS do not fully capture low-frequency and rare variants, but analysis of whole-exome sequencing (WES) reveals rare coding variants, shedding light on novel biological functions for numerous genes and presenting potential avenues for therapeutic development.^19,20^ Due to the evolution of these genetically informed approaches, there has been a growing number of studies systematically assessing strategies to prioritize drug target genes.^21,22^ A recent study examined the efficacy of nominating drug targets from aforementioned genetically informed approaches (GWAS, QTL-GWAS, and Exome) across 30 clinical traits, and further integrated gene network diffusion to prioritize important drug target genes.^21^ Another cross-population meta-analysis study proposed a genomics-driven drug discovery framework comprising three complementary methodologies for gene prioritization, which successfully identified drug targets related to the coagulation cascade for venous thromboembolism.^22^

Here, we applied the core component of developed drug discovery framework^22^ to facilitate identification of therapeutic targets and repurposing opportunities for RA. We performed gene prioritization, overlap enrichment analysis, proteome-wide Mendelian randomization (MR), colocalization, candidate target validation analysis, single-cell RNA-sequencing (scRNA-seq) analysis and population RA incidence association analysis by analyzing the summary statistics from the large-scale RA GWAS, whole-exome sequencing and aptamer-based plasma protein measurements. Our findings provide insight into drug discovery for RA through the systematic integration of multi-layered genetics data.

## Results

### Study overview and summary

The study was designed and conducted as depicted in **Figure1**. First, summary statistics were obtained from the largest large-scale GWAS for RA currently available,^23^ which were derived from 22,350 cases of RA and 74,823 controls in 25 European cohorts (in this study, all analyses were conducted exclusively on individuals of European ancestry, ensuring population homogeneity in genetic and other omics data). We utilized five gene prioritization tools by calculating gene scores or p values to seek for drug candidate genes,^15,24,25,26,27^ after that 65 candidate genes were jointly identified as high priority by four or more tools. Besides, we extracted gene-RA associations based on putative loss of function (pLOF) and deleterious missense variants with MAF <1% from gene burden test results computed on WES data from the UK Biobank,^19^ which top 5% significant genes (940 genes) were likewise treat as candidate genes. To identify opportunities for drug reposition, we then assessed the overlap of candidate genes with targets of existing drugs and quantified an enrichment of the candidate genes in the target of clinical indication categories (i.e., Anatomical Therapeutic Chemical Classification System [ATC]).^22,28^ Out of these candidate genes, 21 were targets of pharmacological agents, which included two approved RA drug targets: *IL6R* targeted by sarilumab and *CD86* targeted by abatacept.

Secondly, we extracted plasma proteome GWAS data derived from the deCODE study, where plasma protein abundances were quantified utilizing 4,907 aptamers across a cohort comprising 35,559 individuals.^17^ Following this, we proceeded with proteome-wide MR analysis^29^ to estimate causal relationships between plasma proteins and RA. Through this analysis, we identified 18 proteins, including tumor necrosis factor (TNF-α) and bromodomain-containing protein 2 (BRD2). Subsequently, we validated these causal relationships using colocalization analysis.^30^ Among the proteins analyzed, hyaluronan and proteoglycan link protein 4 (HPLN4), histone H1x (H1X), and WNT1-inducible signaling pathway protein 1 (WISP1) exhibited high support evidence for colocalization.

Thirdly, we examined whether candidate target genes/proteins interacted with approved RA drug targets in the protein-protein interaction network^31^ and whether the signaling pathways they were significantly enriched in were related to the pathogenesis of RA^32,33^. These validation analyses for utility of candidate target genes/proteins revealed that multiple prioritized genes (for example: colony stimulating factor 1 receptor, *CSF1R*) interactions with 7 targets of current RA medications, and significant enriched signaling pathways of these candidate targets were related to RA development (for example: JAK-STAT signaling pathways).

Finally, to elucidate the biological function of candidate targets for RA, we analyzed scRNA-seq data of joint synovial tissues samples from RA patients.^34^ The scRNA-seq analysis demonstrated that BRD2 widely expressed in joint synovial cells. Then verified whether the most promising target (BRD2) was associated with the onset of RA based on the UKB population cohort.^35^ Each of these parts is described in more detail below.

### Gene prioritization and overlap enrichment in RA relevant medication categories

We employed five gene prioritization tools in parallel, i.e., MAGMA,^24^ DEPICT,^25^ Priority index (Pi),^15^ Polygenic Priority Score (PoPS)^26^ and Transcriptome-wide association studies (TWAS)^27^. Briefly, MAGMA relies on a multiple linear principal components regression framework, integrating linkage disequilibrium (LD) among markers to identify multi-marker effects and ascertain gene significance through p-values.; DEPICT gene prioritization employs a phenotype- and mechanism-agnostic algorithm based on the assumption that truly associated genes share functional annotations, prioritizing genes through scoring, bias adjustment, and false discovery rate estimation steps; Pi represents a scoring system tailored for the drug development landscape within immune-related diseases, and it amalgamates various annotations, encompassing eQTL, chromatin interaction data, and genes associated with immune functions; PoPS leverages the entirety of the polygenic signal while incorporating comprehensive data regarding genes from diverse sources, such as cell-type-specific gene expression and biological pathways, to conduct gene prioritization; TWAS calculates credible sets of causal genes through the utilization of prediction eQTL weights, LD, and GWAS summary statistics. The previous drug discovery framework has demonstrated that the comprehensive integration of five gene prioritization methods can effectively nominate candidate gene sets, thereby enhancing the enrichment of disease-relevant drug-target genes.^22^

The number of genes prioritized by the MAGMA, DEPICT, Pi, PoPS, and TWAS gene prioritization tools is 798, 240, 920, 916 and 3300 (**Figure 2A, Tables S1-S5**) respectively. The intersections of these gene sets determined by different tools are presented in **Table S6**, and 12 genes including *IRF5, PTPN22, IL2RA, IFNGR2, TYK2*, etc are highly prioritized by all five tools. We observed significant enrichment of drug target genes (ORs: 1.65-3.88) in anti-tumor and immune modulator categories (“L” in ATC coding) across the five tools (**Figure 2B, Table S7**). This anti-tumor and immune modulator categories enrichment aligns well with the current mainstream drug categories for treating RA.^36^ Additionally, we examined the overlap and enrichment of these prioritized genes in other ATC codes (such as “M” for the musculoskeletal system); however, we did not identify any significant overlap or enrichment, indirectly suggesting that the concentration of priority genes within the “L” category was not a random occurrence (**Figure 2B**). The omnibus results embracing 65 prioritized genes reach relatively higher enrichment (OR=6.93, *p*=0.00058) (**Table S7**). Notably, this omnibus integration nominated several drug targets including *IL6R* (targeted by tocilizumab/sarilumab, Interleukin-6 receptor alpha subunit inhibitor), which was previously proved to be a successful example of GWAS signals mapped to the target of an indicated pharmacological agent for RA (**Figure 2C, Table S8**).^37^ Likewise, we observed a marginally significant enrichment of drug target categories in anti-tumor and immune modulators (OR=1.58, *p*=0.07) when analyzing the prioritized genes from rare variant burden test results. This finding may be attributed to the fact that not all prioritized genes from the rare variant burden test achieved sufficient statistical significance (all genes-RA associations with *p* > 1×10^-5^) (**Table S9 & S10**). Nevertheless, despite the limitations in statistical significance, this efficiently identified the target gene *CD86*, which corresponds to the clinically approved RA drug abatacept (**Table S10**).^36^

**Figure 1.**
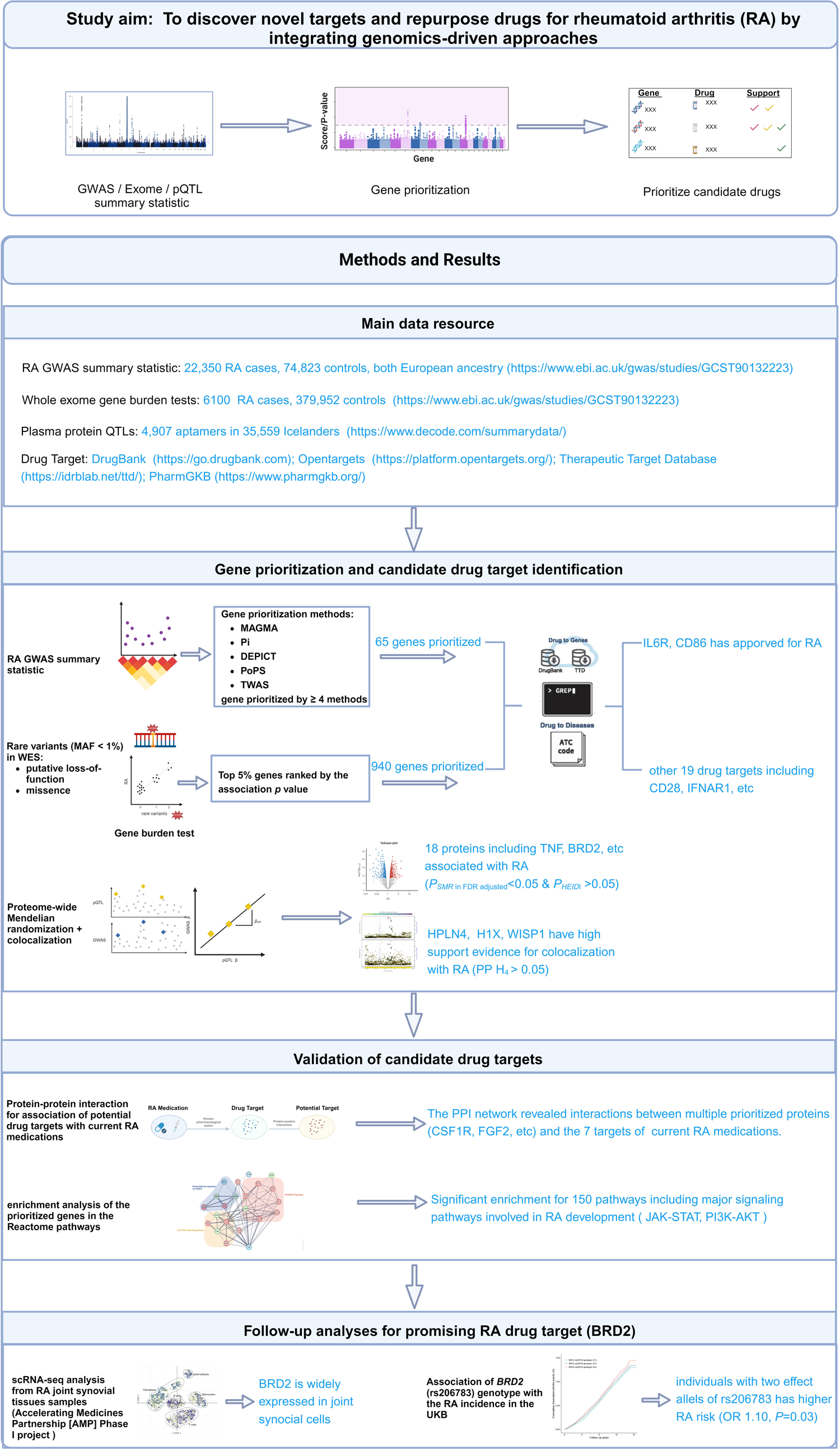
Study overview and summary. We sought therapeutic targets and repurposing opportunities for rheumatoid arthritis by conducting integrative genomics-driven approaches: we conducted gene prioritization based on GWAS and Exome summary statistics, evaluating whether the prioritized genes overlapped with and were enriched in drug targets within the RA-relevant medication category. Concurrently, we identified target proteins through proteome-wide Mendelian randomization and colocalization analysis using pQTL summary statistics. Secondly, we performed protein-protein interaction analysis between the selected candidate drug targets and existing approved RA drugs to explore the potential for drug repurposing. Additionally, we conducted pathway enrichment analysis to assess the biological rationale for their use in RA treatment. Finally, we conducted single-cell sequencing, Phewas, and observational analysis on one of the plausible drug targets, BRD2, to underscore the clinical translational prospects of developing RA therapies linked with it. (This figure created with BioRender.com)

**Figure 2.**
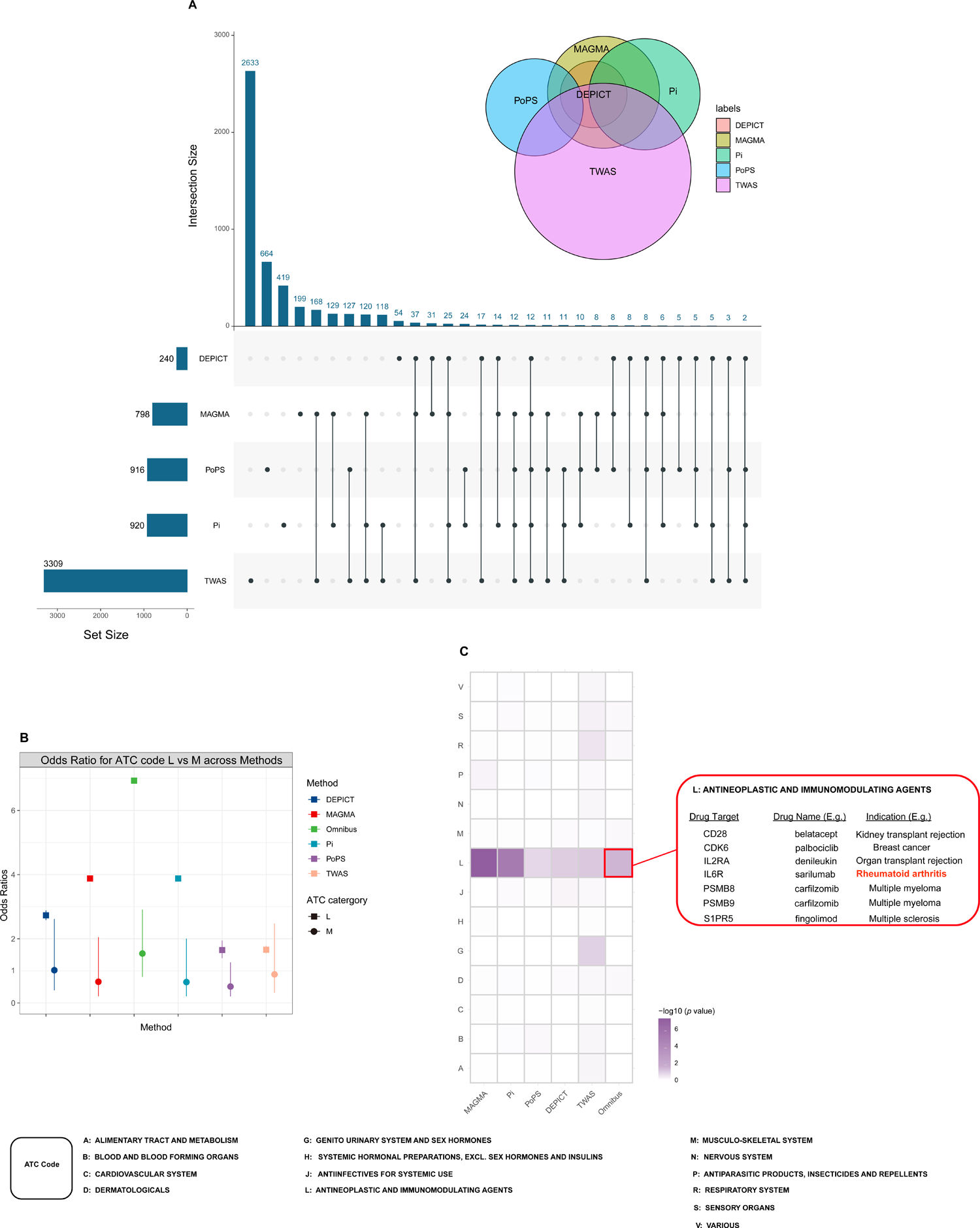
Gene prioritization and overlap enrichment in various medication categories. (A) Set size and intersection size of prioritized genes determined by the five gene prioritization tools (i.e. MAGMA, DEPICT, Pi, PoPS, and TWAS). (B) Overall enrichment of drug-target genes nominated by five gene prioritization tools and their omnibus results. The error bars represent 95% confidence intervals. (C) Enrichments for the five gene prioritization tools and their omnibus results per ATC code. OR, odds ratio.

### Proteome-wide Mendelian randomization analysis identified 18 proteins for RA

Proteome-wide Mendelian randomization uses genetic predictors of protein level to gain insights into the putative causal implications of a multitude of proteins in influencing the risk of various diseases.^38,39^ For our proteome-wide Mendelian randomization analyses, we used *cis*-pQTLs located within a 1 Mb proximity to the transcription start site of the protein-coding gene as genetic instrument, because *cis*-pQTLs are considered to have a higher prior probability of specific biological effects.^40^ Following multiple test corrections, we identified 53 proteins that exhibited significant causality with RA (*P_FDR_* < 0.05, **Table S11**). However, upon further examination using the heterogeneity in dependent instruments (HEIDI) test,^41^ distinguish proteins that were related with RA risk owing to a shared variant rather than genetic linkage, it was revealed that the causal association of 18 out of these 53 proteins with RA was not driven by linkage disequilibrium (*P* _HEIDI_ > 0.05, **Figure 3A & Table S11**). The most prominent causal effect was observed with bromodomain-containing protein 2 (BRD2), where the odds ratio for RA was 26.8 (*P_FDR_=* 3.03×10^-6^) per SD increase in genetically predicted levels (**Table S11**). We performed sensitivity analyses on the identified 18 proteins using either the Wald ratio or the inverse-variance weighted method, found that approximately half of these proteins maintained a significant causal association with RA after applying Bonferroni correction (*P<*0.05/2/18=0.0013, two independent RA GWAS studies and 18 proteins for validation) (**Figure S2**, **Table S12**).

**Figure 3.**
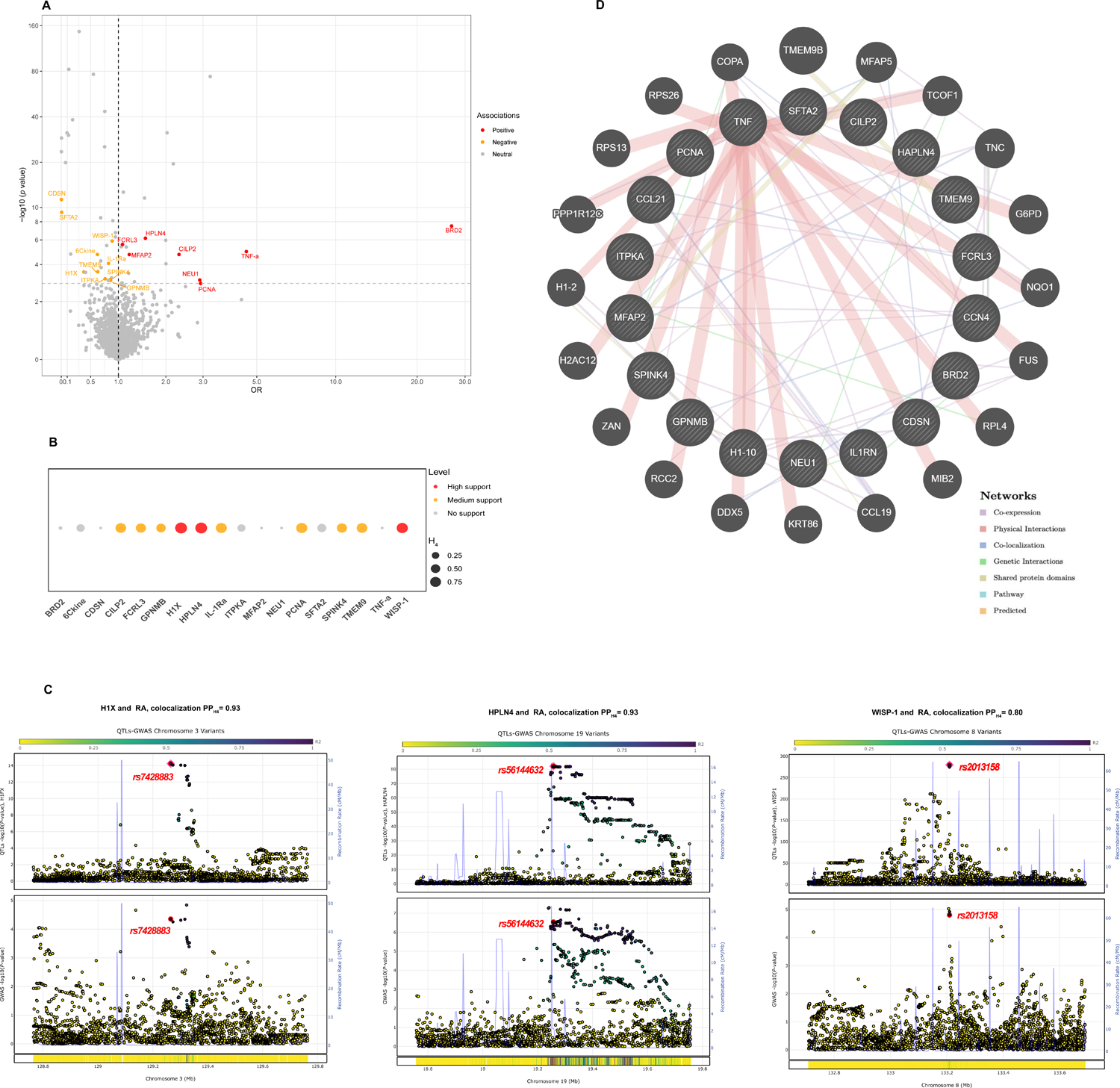
Identification of casual proteins by proteome-wide Mendelian randomization and colocalization analysis. (A) Volcano plot for associations of genetically predicted 1921 plasma proteins levels with RA in MR analysis. Labelled and colour genes refer to MR findings with *FDR*-corrected *P*<0.05 and *P*_HEIDI_ > 0.05. Red genes indicate the positive effect of the plasma proteins on RA; blue genes indicate the negative effect of the plasma proteins on RA. (B) Evidence for colocalization between plasma proteins and RA. Circle size indicates the colocalization posterior probability for H4 and the colour of the circle indicate the classification of the evidence. Evidence support order defined as: *PP_H4_* <0.5 no colocalization support; *PP_H4_* 0.5-0.8 medium colocalization support; *PP_H4_* >0.8 high colocalization support. (C) High support for colocalization with 3 plasma proteins including HPLN4, H1X and WISP1 (Presented sequentially from left to right). (D) Networks of identified causal proteins associated with RA. The network prediction was based on GeneMANIA (http://www.genemania.org).

Among the 18 MR-identified proteins associated with RA, 3 proteins (HPLN4, H1X and WISP1) had high support of colocalization analysis (*PP_H4_* ≥0.8) (**Figure 3B,C & Table S11**). *HAPLN4* (gene corresponding to HPLN4) and *WISP1* (gene corresponding to WISP1) were also known as risk loci for RA.^23^ The co-expression of *H1FX* (gene corresponding to H1X) with *HMGB1*, as illustrated in **Figure S3**, the latter impacts overexpression of signal transduction receptors and abnormal regulation of osteoclastogenesis and bone remodelling possibly linked with progression of RA.^42^ These corresponding genes of 15 identified RA-associated proteins had interaction networks, in particular physical interactions and co-expression (**Figure 3D**).

To verify the prioritized proteins within the framework of drug discovery, we proceeded to collate pharmaceutical compounds from the foremost drug repositories: DrugBank,^43^ Therapeutic Target Database,^44^ PharmGKB,^45^ and the Open Targets Platform.^46^ This yielded 22 drugs relevant to 3 protein-disease associations. (**Table S13**). In addition to TNF-α, a major inflammatory cytokine implicated in the pathogenesis of RA and targeted by several approved monoclonal antibody treatments (such as Infliximab, Etanercept, and Adalimumab),^46^ We also found that literature-based supportive evidence for biologic effects of BRD2 and GPNMB on RA were aligned with putative causality by MR estimates: For instance, Inhibition of BET family proteins (for example: BRD2) suppresses the inflammatory, matrix-degrading, proliferative and chemoattractive properties of rheumatoid arthritis synovial fibroblasts.^47^ Glycoprotein nonmetastatic melanoma protein B (GPNMB) expression in the majority of synovial fibroblasts and tissue-infiltrating monocyte for rheumatoid arthritis and osteoarthritis.^48^

### Validation of the potential of nominated drug targets for treating RA

Integrative approaches have substantial efficiency in screening candidate drug targets: the 21 drug-target genes and 3 actionable proteins in total was nominated (**Figure 4A**). Among the 8 drug targets identified, 3 (*TNF, IL6R*, and *CD86*) were targeted by approved RA medications, while 5 (*CD28, CD19, CSF1R, ESR1*, and *IFNAR1*) were the focus of drugs taken into clinical trials for RA (**Table 1**), thus bolstering the validity of genomics-guided drug discovery and repositioning for RA. Additionally, Furthermore, we conducted a comparison of the prioritized targets — specifically, those involving 65 genes from GWAS, 940 genes from Exome, and 18 proteins from pQTLs — against the druggable genome,^49^ and found that 247 of the 1005 (24.7%) prioritized genes and 14 of the 18 (77.8%) identified proteins overlapped with the druggable genome (**Table S14**).

**Figure 4.**
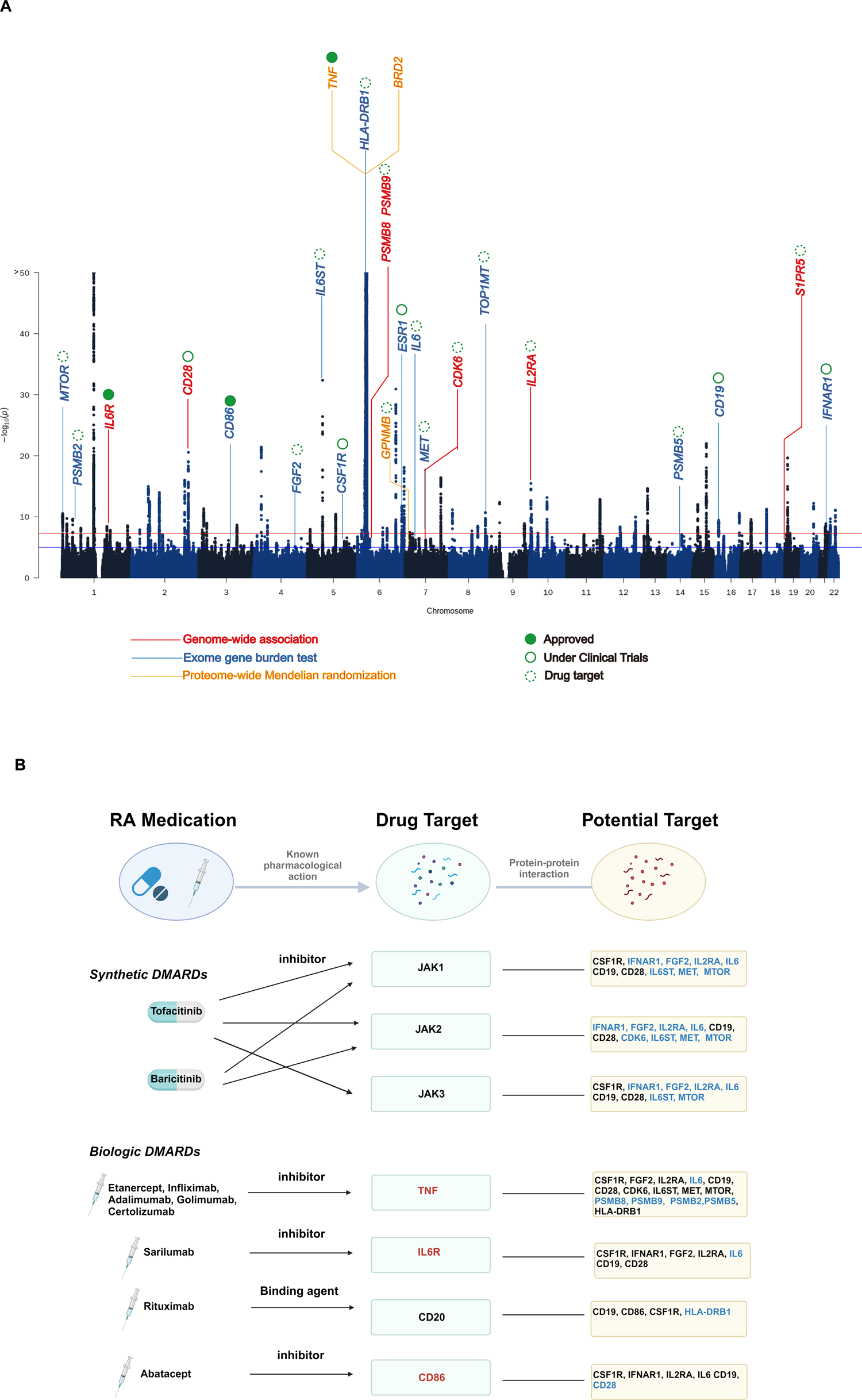
Comprehensive evaluation of drug reposition potentials of nominated candidate drug targets for RA treatment. (A) Candidate target genes nominated for rheumatoid arthritis drug discovery. The colors on the Manhattan chart denote various nomination methods, while the circle type signifies the nominated drug targets at different stages of RA treatment (including approved drugs, those in clinical trials, and targets yet to be clinically tested in RA treatment). (B) Protein-Protein interaction between 6 approved RA medications (7 drug targets) and nominated candidate drug targets. The blue color of nominated candidate drug targets denotes that the type of interaction is known (determined from curated datasets or experiments).

**Table 1.**
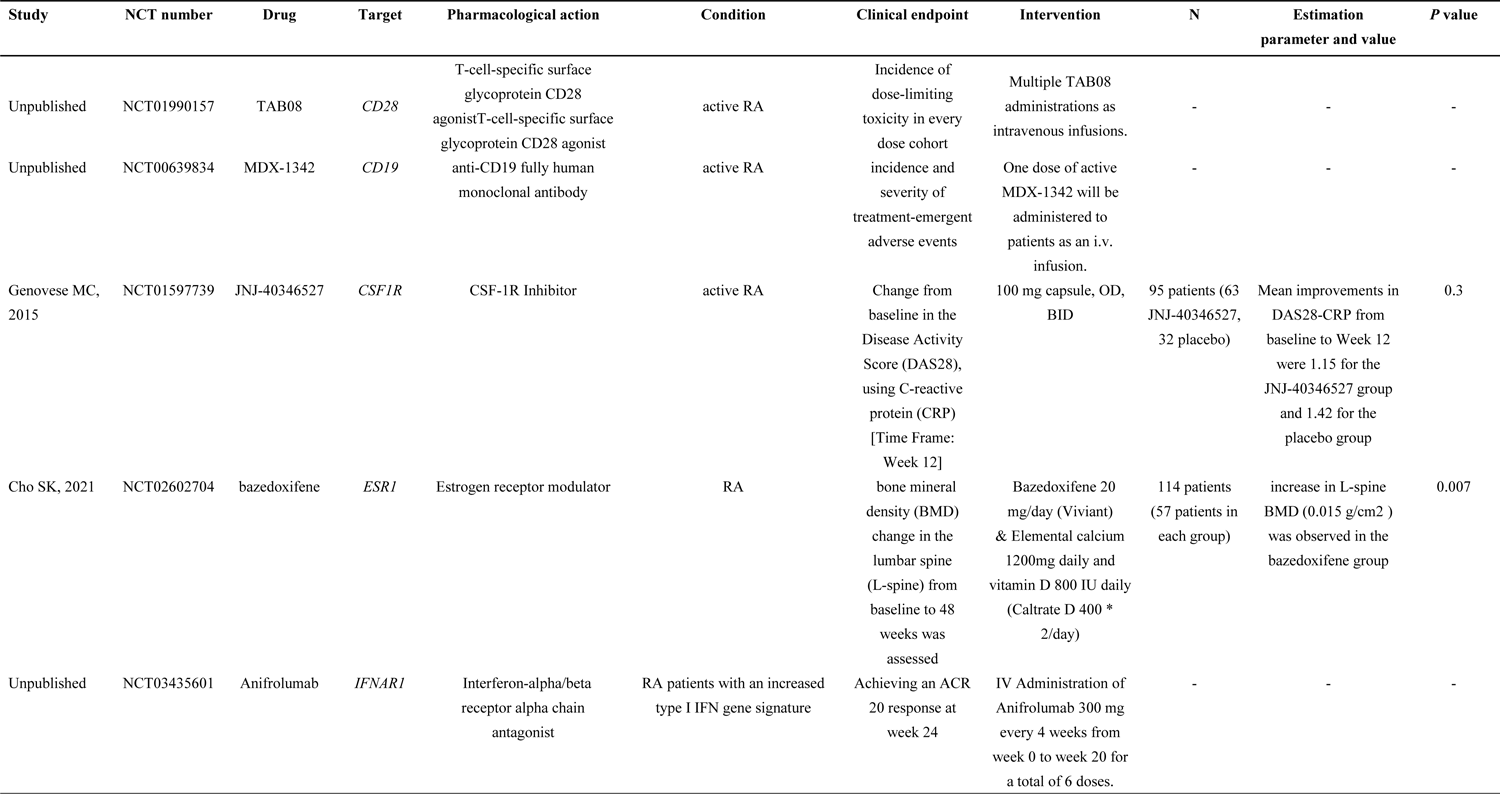
RA-related randomized controlled trails on candidate targets prioritized by integrative genomics-driven approaches.

The PPI network revealed enrich interactions between candidate targets and the seven targets of current RA medications (*TNF, CD86* and *IL6R* were also prioritized targets in our study) (**Figure 4B & Figure S4**). *JAKs-MTOR, MET, CDK6, IL2R, FGF2* and *IFNAR1* were determined to have known interactions. For example, *JAK1* can be phosphorylated by by four cytokine-receptor families such as *IFNAR1* and *IL-2R*,^50^ which have been validated as pathogenic pathways in RA.

We conducted further evaluation of the pathway enrichment of the candidate targets and observed significant enrichment for 150 pathways from Reactome (**Table S15**), as well as 20 from KEGG (**Figure S5**). Relevant signaling pathways implicated in the pathogenesis of RA were validated, including the JAK-STAT and PI3K/AKT signaling pathways. Particularly noteworthy was the transcriptional regulation by RUNX3 signaling pathway (*P_FDR_* = 3.26×10^−5^), which was nominated by integrating three drug-discovery approaches (**Figure 5**). This pathway included five genes separately prioritized by the three approaches: *PSMB8* and *PSMB9* by the omnibus gene prioritization approach, *BRD2* by proteome-wide MR, and *PSMB2* and *PSMB5* by the rare variant burden tests from Exome. RUNX3 signaling pathway might play roles on the mechanisms of T cell activation in RA.^51^ Our finding demonstrates that the three approaches synergistically prioritized therapeutic targets for RA. We also observed *BRD2, PSMB2, PSMB5, PSMB8* and *PSMB9* were almost moderate to high expression in most human tissues (**Figure S6**).

**Figure 5.**
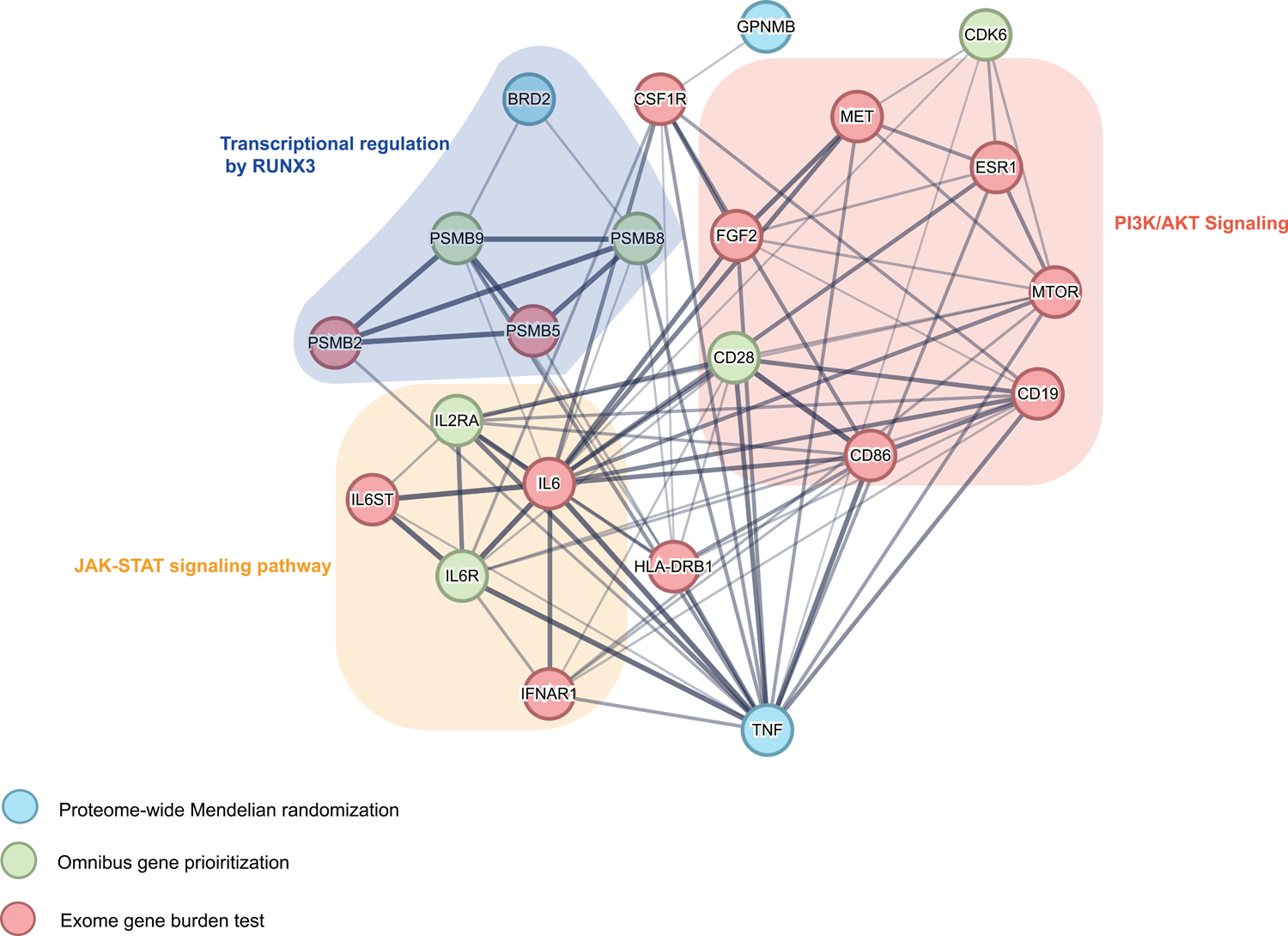
The three drug discovery approaches complementarily prioritized drug targets for RA. The genes prioritized for RA by the three drug discovery approaches were connected to each other if their protein-protein interaction scores were larger than 0.4. Line thickness represents the strength of protein-protein interaction.

### Follow-up analyses for the putatively therapeutic target-BRD2

To explore whether the candidate targets had any cell type-specific enrichment in joint synovial tissues, we further performed single cell-type expression analysis using data from 5,265 scRNA-seq profiles of 36 RA patients (Single-Cell Portal of the Broad Institute, AMP Phase 1 study). Our observations indicated that BRD2 exhibits a notably widespread expression across various cell types in joint synovial tissues, especially when compared to other candidate targets (**Figure 6A**). The expression values of *BRD2* in each cell, positioned in a t-Distributed Stochastic Neighbor Embedding (tSNE^52^) plot, and relatively higher expression values of *BRD2* were found in B cells and T cells (**Figure 6B**). These cells correspond to 18 clusters within 5 cell types, namely fibroblasts, B cells, monocytes, plasmablasts, and T cells (**Figure 6C**). Specifically, we found that *BRD2* was significantly expressed in 2 types of B cells (naïve IGHD+CD27- and autoimmune-associated B cells), plasmablast, 3 types of fibroblasts (HLA-DRAhi sublining, DKK3+ sublining and CD55+ lining fibroblasts), NUPR1+ monocytes, 4 types of T cells (CCR7+ and three CD8+ clusters: GZMK+, GNLY+GZMB+ cytotoxic lymphocytes, and GZMK+GZMB+ T cells) (permutation test *P* < 0.001; **Figure 6D**). We also found *BRD2* differential expression between rheumatoid arthritis patients (n=315) and healthy control (n=315) in CD14dim.CD16+ cell from Rheumatoid Arthritis Bioinformatics Center^53^ (RABC, dataset id: RABC18) (**Figure S7**).

**Figure 6.**
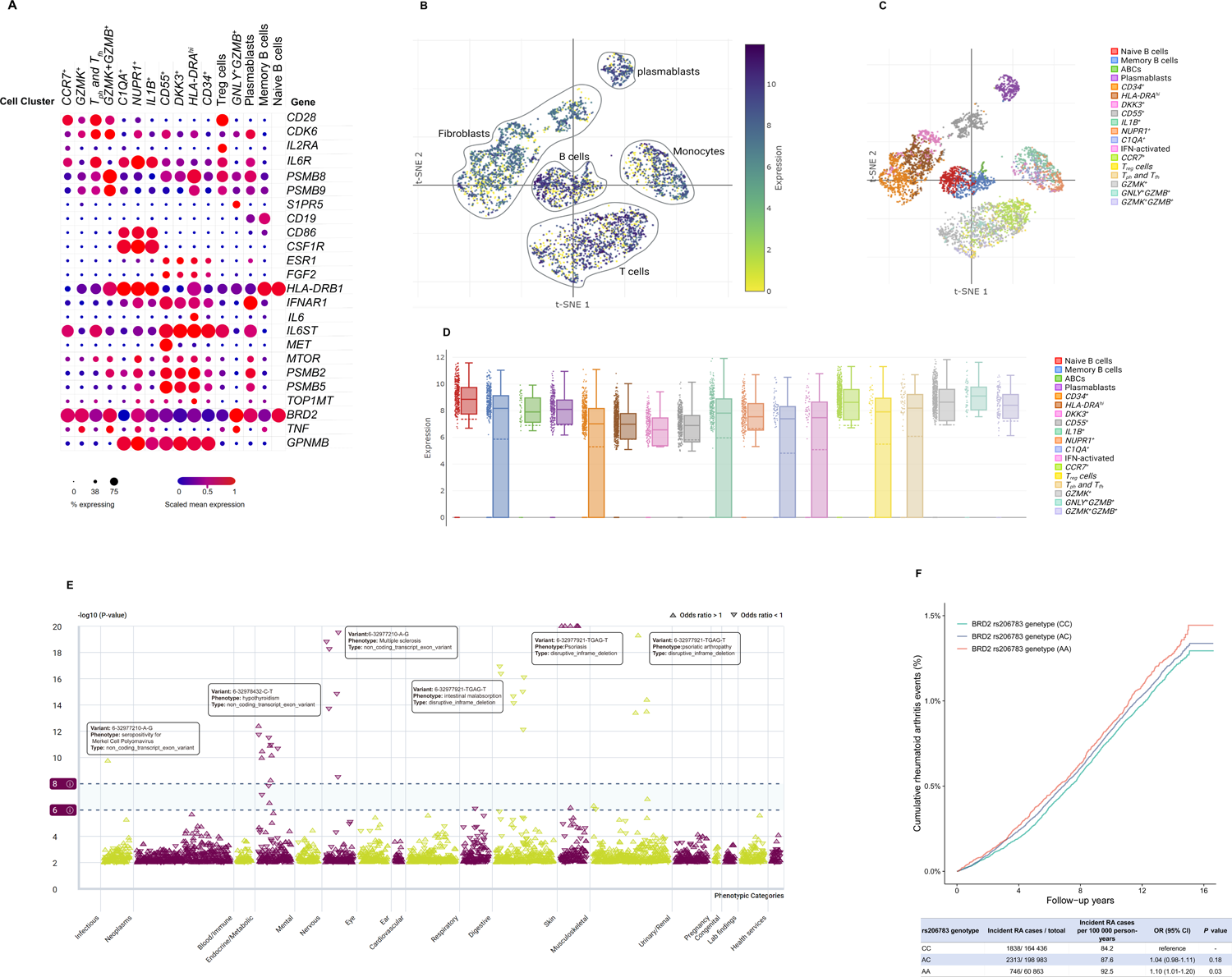
Multiple layers of evidence support the clinical efficacy of putative drug targets in RA treatment. (A) Candidate drug target genes expression levels of each cell type at single-cell resolution in the joint synovial tissues of 36 RA patients. (B) BRD2 expression levels of each cell type (C) 18 annotated cell types clusters across 5,265 cells from all cell types in the joint synovial tissues. (D) BRD2 expression status in 5,265 scRNA-seq profiles, including 1,142 B cells, 1,844 fibroblasts, 750 monocytes, and 1,529 T cells. BRD2 expression levels of 18 cell types are shown in the box plot. In each box, the horizontal solid line represents the median of the expression level, and the horizontal dashed line represents the average value. Each box extends from the 25th percentile of each group to the 75th percentile. Whiskers extend 1.5 times the interquartile distance from the top and bottom of the box. (E) Manhattan plot for PheWAS of BRD2 at variant level. 35 variant-trait pairs reach the significant genome-wide association threshold (*P* < 5 × 10 ^−8^), 53 variant-trait pairs reach the suggestive threshold (*P* < 5 × 10 ^−6^). Upward triangle represents positive association, downward represents negative association. (F) Cumulative risk of RA incidence between differen rs206783 genetype (AA, AC, CC) in UK Biobank population (424,282 individuals, 4897 incident RA cases). rs206783 is the index *cis*-pQTL of BRD2.

Given that *BRD2* was identified as one of the highest confidence targets supported by diverse approaches, we sought to investigate whether *BRD2* might have potentially beneficial or deleterious effects on traits beyond RA. To accomplish this, we screened the results from a phenome-wide association study (PheWAS) performed on the AstraZeneca PheWAS Portal.^54^ While *BRD2* itself was not significantly associated with any other traits at the gene level (**Figure S8**), an indel variant (6-32977921-TGAG-T) of *BRD2* was found to be positively associated with psoriasis and psoriatic arthropathy, which the odds ratios (ORs) and p-values for these associations were 1.95 (9.12 × 10 ^−122^) and 2.50 (6.36 × 10 ^−36^), respectively.

Furthermore, a missense variant (6-32974662-A-G) of *BRD2* was observed to be associated with multiple sclerosis, with an OR of 9.30 (P = 0.0052) (**Figure 6E & Table S16**). These evidence suggesting that inhibition of bromodomain proteins have potentiality in drug reposition for rheumatic diseases.^55^

To triangulate the evidence regarding the effect of *BRD2* on RA, we further evaluated whether rs206783 (the top *cis*-pQTL of *BRD2*) was associated with RA incidence in the general population, such as in the UK Biobank dataset. Our analysis revealed that individuals with the genotype (AA) of rs206783 exhibited a slightly higher risk of RA, with an odds ratio (OR) of 1.10 (95% CI: 1.01-1.20), and a p-value of 0.03, when compared to those with the genotype (CC) (**Figure 6F**). The significant relevance between rs206783 and RA was reaffirmed in the FinnGen study using RA GWAS summary statistics rather than population observational data (**Table S17**).

## Discussion

In the present study, we integrated three genomics-driven approaches (GWAS, QTL-GWAS, and Exome) to prioritize drug target genes/proteins. We further evaluated their biological and clinical relevance with RA to ascertain their utility in discovering candidate drug targets. GWAS and Exome analysis complement each other in prioritizing genes for drug targeting when considering overlap with drug target in RA-relevant medication codes (primarily anti-tumor and immune modulator categories). Of note, both approaches corroborated *IL6R* (targeted by approved RA drug sarilumab) and *CD28* as candidate drug targets for RA. Whereas currently, only a relatively few clinical trials have proved the abilities of remaining candidate targets such as *IFNAR1* antagonist [Anifrolumab] and *CSF1R* Inhibitor [JNJ-40346527] in achieving remission of RA. Furthermore, proteme-wide MR improved drug development yield by revealing causality of 15 proteins with RA beyond gene prioritization. The two causal proteins (BRD2 and GPNMB) have the potential as therapeutic targets for RA as indicated in the supporting literatures.^47,48^ The subsequent single-cell RNA sequencing analysis revealed high expression of *BRD2* in immune cells and fibroblasts, indicating its possible role in inflammation and fibrogenesis, thereby triggering the development of RA. Finally, we assessed the distinct effects of *BRD2* on wide-range traits through phenome-wide association studies (PheWAS) and found its top *cis*-pQTL (rs206783) significant association with RA incidence at the population level.

The gene priority determined by the Exome method mostly differs from that determined by the GWAS method, a distinction also reflected in the identified drug targets. The odds ratio of drug target enrichment in the anti-tumor and immune modulator medicine categories for the Exome method was 1.58 (*P*=0.068), which was significantly lower than that of the GWAS method (OR 5.19, *P*=0.002). This difference may be attributed to two situations: Firstly, the Exome method exclusively focuses on rare variations with a minor allele frequency (MAF) less than 1% in the WES data. The gene score is derived from the gene load test of functional loss and missense variation, which may lead to relatively low power, a trend consistent with previous studies comparing gene prioritization methods.^21^ Secondly, gene prioritization based on the GWAS method is an omnibus strategy (Contains five gene prioritization methods with respective advantages), and one of its components, PI, could lead to a considerably higher enrichment in immune drugs, as consistently reflected in our results.^15^

The confirmation of *CD28* as a candidate drug target by both GWAS and exome analysis is notable. *CD28* superagonists (CD28SA), exemplified by TGN1412, is a CD28-specific monoclonal antibody known to activate T cells without substantial TCR involvement.^15^ However, early-phase I clinical trials of TGN1412 encountered severe cytokine storms, prompting termination of development. Subsequent investigations suggested that appropriate dosages, such as those used for TAB08 (another *CD28* superagonists), could selectively activate regulatory T cells without inducing significant release of pro-inflammatory cells (Clinical trials identifie: NCT01885624). Phase Ib trials in RA patients demonstrated acceptable adverse events level associated with TAB08 (Clinical trials identifier: NCT01990157). Future phase II trials are needed to further establish TAB08’s capacity to activate regulatory T cells for immune diseases. Colony-stimulating factor 1 receptor (*CSF1R*) emerged as another promising drug candidate target in our study. *CSF1R* plays a crucial role in modulating monocyte proliferation, migration, and activation, all of which are considered significant factors in the pathogenesis of RA. Increased expression of *CSF1R* has been observed in the synovium of RA patients,and blockade of *CSF1R* has demonstrated protective effects against bone and cartilage destruction in mouse models.^57^ However, the results of a phase IIa parallel-group study (Clinical trials identifier: NCT01597739) investigating the oral *CSF1R* inhibitor JNJ-40346527 in patients with active rheumatoid arthritis did not show a significant improvement in disease activity or inflammatory levels.^58^ These studies collectively indicate that there remains a substantial gap between genetic-supported drug candidates and their actual clinical applications, underscoring the need for further in-depth research and verification.

Mendelian randomization contribute to genetic drug target validation, which utilized instrument variable to proxy drug target and estimated causal relation with diseases.^38,39^ Considering that proteins are typically the direct targets of drugs in most cases (over 90% of drug targets are proteins), protein exposure is usually the preferred choice for drug target MR. A prior investigation assessed the causal effects of inflammation-related proteins across immune-mediated disorders, elucidating the directionally disparate impact of CD40 on the susceptibility to RA compared to other autoimmune conditions such as multiple sclerosis and inflammatory bowel disease.^15^ While we were unable to replicate the causal association between CD40 and RA in proteome-wide MR analysis due to the unavailability of CD40 pQTLs for present MR testing, our study did demonstrate that four out of the five gene prioritization methods based on GWAS jointly confirmed its prioritization. Furthermore, in situations where genetic instrument variables for related proteins are unavailable, upstream markers such as mRNA or downstream markers can also serve as indirect confirmations of alternative exposure.^59^ A recent study investigated the association between cis-expression quantitative trait loci (*cis*-eQTL) and RA risk using MR and colocalization analysis as the primary study design. The study identified *CCR6, HLA-DPA1, HLA-DRB1, IFNGR2, C5, ATP2A1, and FEN1* as potential candidate drug target genes, and candidate drug prediction and molecular docking were utilized for further verification.^60^ We study used proteome-wide MR analysis based on *cis*-pQTL and obtained a list of candidate drug targets that were completely different from this study, this discrepancy indicated the MR estimation of tissue-specific drug targets through eQTL weighted analysis may not always align with evidence from similar blood-based pQTL analysis or drug trials.^59^ While our study and existing research did not yield shared determined actionable subjects through drug target MR, these findings provide valuable insights into drug target prioritization and medicinal potential from different perspectives. Among the 15 causal proteins identified by our study, three of them (HPLN4, H1X, and WISP1) were verified to have colocalization support with RA. However, little is known about the potential role of these three proteins in the pathogenesis of RA. H1X may contribute to the induction and persistence of complement activation and arthritis in RA, as it is one of many citrullinated peptides found in the synovial fluid of RA patients, which are targets of the anti-citrullinated protein antibody response.^61^ Genetic polymorphisms in *WISP1* have been associated with RA susceptibility,^23^ WISP1 plays a significant role in regulating Wnt signaling in bone homeostasis,^62^ which might be linked to the progression of RA. *HAPLN4* has been identified as a drug-targeting gene for schizophrenia and bipolar disorder.^63^ Additionally, existent evidence have shown consistent effect directions of multiple genetic variants between schizophrenia-RA and bipolar disorder-RA pairs.^64^ These findings suggest that *HAPLN4* may serve as a therapeutic target not only in psychiatric disorders but also in immune disorders like RA. This indicates potential shared mechanisms or pathways underlying these seemingly distinct indications, opening avenues for further exploration into their treatment.

The series of results from our current study suggest promising clinical prospects for BRD2 in the treatment of RA. BRD2 is belong to the bromodomain and extra-terminal (BET) family member proteins. BRD2, BRD3, and the most studied BET protein BRD4 are ubiquitously expressed in mammalian cells and tissue types.^65^ A recent investigation has illustrated that targeted suppression of the second bromodomains within BET proteins (referred to as iBET-BD2) efficiently attenuated the synthesis of pivotal pro-inflammatory mediators, notably Th17 cytokines, within a co-culture system comprising B and T cells. Remarkably, this inhibition exerted no discernible influence on the proliferative capacity of the cells, yet it notably diminished the secretion of effector cytokines, including IFNγ, IL-17A, and IL-22. Subsequently, the study tested an optimized iBET-BD2 compound (GSK620), which showed a significant dose-dependent inhibition of both arthritic score and IgG1 production in response to immunization in a collagen-induced arthritis model in rats.^66^ Additionally, BET inhibitors influence the inflammatory responses of various immune cells, including B cells, macrophages, synovial fibroblasts, and chondrocytes, indicating a broad role of BRD proteins in autoimmune disorders.^54^ These finding aligns with the anticipated effects of inhibitors for BRD2 on RA. The extensive and high expression of BRD2 observed in B cells, T cells, and fibroblasts in joint synovial tissues, as identified in our current study, suggests a potential role for BRD2 inhibition in modulating the inflammatory response associated with RA.

In summary, our study has facilitated the in *silico* screening of abundant drugs and targets with supporting evidence. It underscores that thorough evaluation of genetic evidence offers indispensable potential in advancing clinical therapy for RA.

### Limitations of the study

Several limitations should be considered. First, the primary focus of this study lies in examining the overlap and enrichment of prioritized genes with the drug targets in anti-tumor as well as immunosuppressive medications, potentially overlooking effective targets within alternative categories. Second, despite encompassing a wide array of blood proteins in this study, there is a possibility that we have inadvertently neglected significant proteins lacking genetic instruments. Moreover, besides proteins, other intermediary molecular quantitative traits (eQTL, mQTL, sQTL) may offer further insights. Third, subtle difference in population structure between pQTL (Only Iceland population) and GWAS (Multiple European population). Fourth, for the observational analysis and verification of the association between BRD2 and RA, we did not directly utilize the standard expression of BRD2 (as measured by the UKB through the olink platform). This decision stemmed from the relatively small number of incident RA cases within the population with available standard protein expression data for BRD2, potentially resulting in insufficient statistical power. Finally, while we have substantiated some prospective targets by scrutinizing evidence derived from biological efficacy or clinical trials, it is imperative that direct in *vivo* or in *vitro* experimentation be undertaken as the subsequent phase for the prioritized targets identified in this investigation.

## Data Availability

The RA GWAS summary statistics and whole exome gene burden tests are publicly available at https://www.ebi.ac.uk/gwas/summary-statistics. Plasma pQTL summary statistics are obtained from deCODE study at https://www.decode.com/summarydata/. The genetic and phenotypic UK Biobank data are available on application to the UK Biobank to any researcher worldwide (www.ukbiobank.ac.uk).

The genomics-driven drug discovery analysis was conducted using the following publicly available tools: MAGMA (https://ctg.cncr.nl/software/magma), DEPICT (https://data.broadinstitute.org/mpg/depict/), PoPS (https://github.com/FinucaneLab/pops), FOCUS (https://github.com/bogdanlab/focus), GREP (https://github.com/saorisakaue/GREP), SMR (https://yanglab.westlake.edu.cn/software/smr/), ezQTL (https://analysistools.cancer.gov/ezqtl), GeneMANIA (https://genemania.org/), FUMA (https://fuma.ctglab.nl/), PheWAS (https://azphewas.com/) and the “Pi”, “TwosampleMR” and “coloc” R packages.

## Acknowledgments

We acknowledge investigators and participants in the UK Biobank study, the deCODE project and cited genome-wide association studies for sharing data. The authors also thank Nanjing Medical University Prof. Xianyong Yin for guide on statistical methodology. This study was supported by Research Funds of Joint Research Center for Occupational Medicine and Health of IHM(OMH-2023-01;OMH-2023-08) and Anhui Province clinical medical research transformation project (NO.202304295107020041; NO.202304295107020048.

## Author contributions

Jie Zhang conceived and designed the study. Jie Zhang and Xinyu Fang performed statistical analyses. Jie Zhang drafted the manuscript. Dongqing Ye supervised all statistical analyses. All the authors contributed by providing advice on interpretation of results. All authors read, approved, and provided feedback on the final manuscript.

## STAR★Methods

### Key resources table

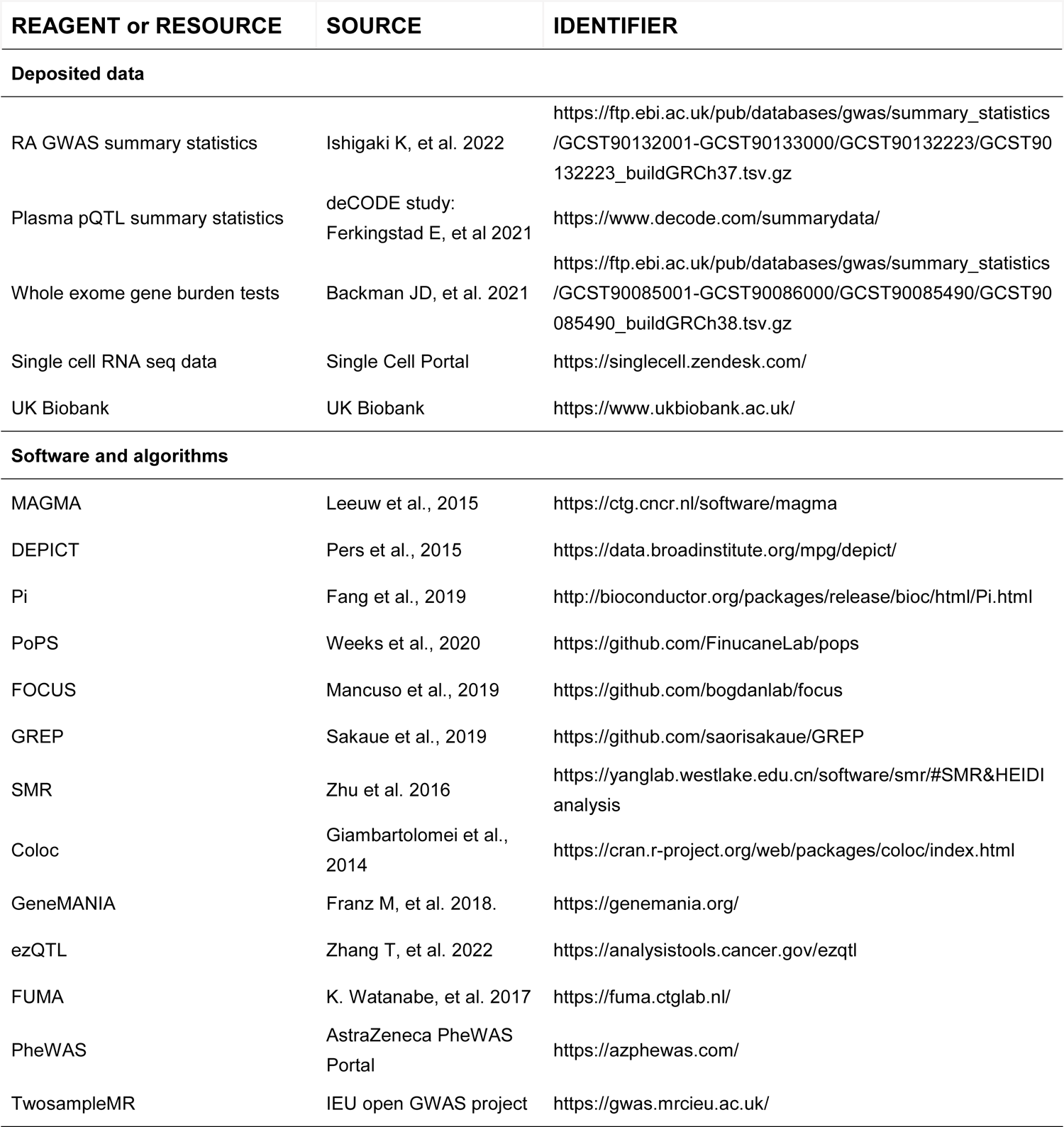

#### Resource availability Lead contact

Further information and requests for resources should be directed to and will be fulfilled by the lead contact, Dongqing Ye.

#### Materials availability

This study did not generate new unique reagents.

## Method details

### Genetic summary statistics resources

GWAS summary statistics for rheumatoid arthritis (RA) were obtained from the Ishigaki K, et al’s study^23^ with 22,350 cases of RA and 74,823 controls of European ancestry. The summary statistics data can be accessed via the GWAS Catalog using the access ID: GCST90132223.

We utilized gene burden test outcomes derived from whole-exome sequencing (WES) data obtained from the UK Biobank. Gene-RA associations were extracted, focusing on putative loss-of-function (pLOF) and deleterious missense variants with a minor allele frequency (MAF) below 1%. These associations were available for approximately 18,800 genes, ranked by association p-value, and retrieved using the provided Ensembl identifier.

Ferkingstad et al., 4719 proteins measured in 35,559 Icelanders, were employed to extract summary statistics of genetic associations with plasma proteins.^17^ We extracted the *cis*-pQTL variants under multiple-testing significance threshold (determined using Bonferroni correction for all 27.2 million tested variants, giving a threshold of 0.05/27,200,000=1.8 × 10 ^−9^), and *cis*-pQTL was defined as a SNP residing within 1 megabase (Mb) upstream or downstream of the transcription start site (TSS) of the corresponding protein-coding gene.

### Gene prioritization

We used five distinct tools for gene prioritization based on RA GWAS summary statistics, namely MAGMA^24^, DEPICT^25^, Priority Index (Pi)^15^, Polygenic Priority Score (PoPS)^26^, and Transcriptome-wide Association Studies (TWAS)^27^. We analyzed with default settings unless explicitly specified otherwise.

MAGMA is a gene analysis method that accounts for LD structure, which projects the SNP matrix for a gene onto its principal components (PC), obtained PCs without small eigenvalues as predictors to examine association between genes and phenotype in the linear regression model. For MAGMA,^24^ “gene-model snp-wise = mean” option was set. DEPICT operates by taking a collection of trait-associated SNPs as input, leveraging them to detect independently associated loci, which may encompass multiple genes. DEPICT predicts the biological functions of genes across a broad spectrum of biological annotations, comprising 14,461 gene sets. Subsequently, basing on this information of gene sets, DEPICT identifies reconstituted gene sets that exhibit enrichment for genes within the associated loci, and prioritizes genes within associated loci by identifying genes from different loci that share similar predicted functions. We selected these variants with relaxed p-values < 1.0×10^− 5^ as input for DEPICT.^25^ Pi is primarily constituted by a gene-predictor matrix that integrates genomic and annotation predictors, aiming to prioritize approximately 15,000 genes for a given trait. Initially, Pi identifies “Seed genes” by leveraging scores from genomic predictors to ascertain functional linkage to the input disease-associated genetic variant, considering factors such as proximity, conformation, and expression. Subsequently, scores from annotation predictors related to immune function, phenotype, or disease are exclusively applied to these seed genes. Non-seed genes are determined based on knowledge of network connectivity. The predictor matrix generates a numerical Pi prioritization rating ranging from 0 to 5, along with corresponding rankings, while affinity scores ensure comparability across different predictors for both seed and non-seed genes. Following the methodology of the original study,^15^ we utilized lead variants with a significance threshold of p < 5.0×10^− 8^ as input, eQTL in the peripheral blood and immune cells, chromatin interaction in immune cells, topologically associating domain boundary in the GM12878 cell line, and the STRING^31^ protein-protein interaction network (PPI), with a high confidence score. PoPS is a tool that attributes high scores to genes exhibiting features akin to those with robust gene-level associations. It ran under the assumption that genes located proximally to associated SNPs and sharing analogous biological annotations are more likely to be causally related. PoPS utilizes gene-level associations derived from GWAS summary statistics to ascertain collective polygenic enrichments of gene features sourced from cell-type-specific gene expression, biological pathways, and PPI. In order to nominate potential causal genes, PoPS assigns a priority score to each protein-coding gene based on these enrichments.^26^ In this context, we opted to employ top-ranked genes rather than pinpointed genes, thereby incorporating multiple genes per locus for drug discovery purposes, as the conduction done by previous study.^22^ We conducted FOCUS^27^ to disentangle the causal and tagging gene-trait associations at a TWAS-significant region, which FOCUS serves as a fine-mapping tool, estimates credible sets of causal genes by integrating prediction eQTL weights, LD, and GWAS summary statistics. We iteratively executed FOCUS across 44 GTEx v7 tissues, and calculated the median of their posterior inclusion probabilities (PIP).

We prioritized genes according to predefined thresholds as follows: for MAGMA, genes were prioritized if they exhibited a FDR < 0.05; for Pi and PoPS, the top 5% of genes were selected based on descending gene scores; for DEPICT, a more inclusive threshold of FDR < 0.2 was applied, as DEPICT calculates p-values exclusively for genes within pre-featured target loci by default; and finally, genes were prioritized from TWAS results if they demonstrated a PIP > 0.1.

### Proteome-wide Mendelian randomization (MR) analysis

Summary-data-based Mendelian randomization (SMR) analysis was conducted to estimated the causal associations between proteins and RA.^67^ We used the following settings: --maf 0.01, --thread-num 10, --diff-freq 1. To control for genome-wide type I error, we conducted FDR correction.

To distinguish proteins that were related with RA risk owing to a shared variant rather than genetic linkage, we further examine using the heterogeneity in dependent instruments (HEIDI) test.^41^ The *p* value of the HEIDI test > 0.05 indicated that the association of protein and RA was not driven by linkage disequilibrium.

The“TwoSampleMR”RA package was employed to perform Replication MR analysis on 18 proteins identified in SMR analysis. For any proteins with only one instrument, the Wald ratio method was used to estimate the log odds change in RA risk for per standard deviation (SD) increment of circulating protein levels as proxied by the instrumental variables. The inverse-variance weighted (IVW) method was used to obtain the MR effects estimates for proteins with more than one instrument. The heterogeneity test was performed to assess the heterogeneity of the genetic instruments based on the Q statistic. We also performed additional analyses including MR-Egger to account for horizontal pleiotropy. Replication MR analysis was further performed for the identified proteins based on RA GWAS summary data from Sakaue, et al (2021) (ebi-a-GCST90038685) and Okada, et al (2014) (ieu-a-832), respectively. Bonferroni correction was used for multiple testing correction, with *P<*0.0013 (0.05/2/18, two independent RA GWAS studies and 18 proteins for validation) as the significance level.

### Colocalization analysis

We employed a Bayesian colocalization approach to identify shared genetic variants between RA GWAS signals and *cis*-pQTL using “coloc” R package.^30^ We used the default coloc priors for Bayesian colocalization analysis, with p1=1×10^−4^ (prior probability a SNP is associated with protein), p2=1×10^−4^ (prior probability a SNP is associated with RA), and p12=1×10^−5^ (prior probability a SNP is associated with both protein and RA). For each protein (establishing causality with RA determined by our SMR analysis), we extracted the region located within 1 Mb around the cis-pQTL. Five posterior probabilities (PPs) were calculated for the colocalization analysis: PP_0_ (null model of no causal genetic variant); PP_1_ (probability that causal variants only associated with protein); PP_2_ (probability that causal variants only associated with RA); PP_3_ (probability that RA and proteins causal variants are distinct), and PP_4_ (probability that RA and proteins share same causal variants). The genes were defined as high support for colocalization with cut-off of PP_4_ ≥ 0.8. The visualization of colocalization is achieved through ezQTL website based tool.^68^

### Function and network prediction

We utilized GeneMANIA to predict the biological network and functions of cis-genes associated with RA-related proteins. Further details regarding the datasets incorporated into GeneMANIA can be found elsewhere.^69^

To explore the potential interactions between identified candidate target and current RA medication targets, a PPI network was constructed using the STRING database (https://string-db.org/), we retained the protein-protein pairs with interaction scores larger than 0.4.

### Clinical evidence evaluation

We undertook a systematic review focusing on RA-related clinical trials targeting candidate targets. Our search encompassed studies across three databases: MEDLINE, EMBASE, and the clinical trials registration database, with publications up to February 30th, 2023. Trials that were not randomized controlled trials (RCTs) or not conducted on human subjects were excluded from our analysis. Key information, including the first author, year of study, National Clinical Trial number, patient characteristics, sample size, intervention, trial phase, trial status, assessment of efficacy, and adverse effects, were systematically extracted from eligible studies.

### Single cell-type expression analysis

To disentangle the BRD2 expression pattern in the joint synovial tissues of RA patients, we obtained 5,265 scRNA-seq profiles of 36 RA patients from Zhang, F. et al study.^34^ from the Single-Cell Portal of the Broad Institute (https://singlecell.broadinstitute.org/single_cell/) (accession ID SCP279).

We reanalyzed the data, focusing on BRD2 expression status. We analyzed the gene expression matrix and the associated metadata using R v.4.1.2 and “Seurat” R package (https://satijalab.org/seurat/). To cluster the cells, we adopted the clustering annotation from the original study^34^. To test whether BRD2 expression was enriched in specific cell types, we calculated the proportion of BRD2-expression cells in this cell type. Subsequently, we permuted the cell type labels 1,000 times and obtained the frequency (permutation *P* value) of the same cell type containing the same or a larger proportion of BRD2-expression cells. Additionally, we compared the BRD2 expression level between a target cell type and the other cell types using Wilcoxon rank-sum test.

### UK Biobank genotyping data and quality control

The genotypes of the UK Biobank participants were assayed using either of two genotyping arrays, the Affymetrix UK BiLEVE Axiom (approximately 50,000 participants) or Affymetrix UKBiobank Axiom genotyping array (approximately 450,000 participants), there are 805,426 markers inthe released genotype data. These arrays were augmented by imputation of approximately 90 milliongenetic variants from the Haplotype Reference Consortium and the UK 10K projects. Details of the array design, sample processing, and stringent quality control have been previously described.^35^ we further excluded participants with sex mismatch, heterozygosity rate outliers, and missing genotypes, with excess relatives and non-European ancestry (non self-report British white or Caucasians ascertained by genetic principal components analysis), as well as prevalent RA paticipants (ICD-9 codes: 714; ICD-10 codes: M05 and M06; Self report RA:1464), resulting in a final cohort of 424,282 individuals (including 4897 incident RA cases)

### Quantification and statistical analysis

#### Enrichment analysis in medication categories

We used the prioritized gene list as the primary input to conduct a sequence of Fisher’s exact tests concerning ATC codes, aiming to assess the enrichment of drug-target genes within specific codes. The drug-target database utilized in this analysis was sourced from GREP,^28^ curated from two prominent drug repositories, Drug Bank and TTD. The Anatomical Therapeutic Chemical (ATC) classification system serves as a pivotal framework for categorizing drugs, where approved medications can belong to multiple ATC codes. We selected the ATC code L as RA relevant medication category: anti-tumor and immune modulator, characterized by the highest count of approved drugs tailored for RA treatment in this ATC code medication category.

#### Pathway enrichment analysis

We conducted one-sided hypergeometric tests utilizing the “ReactomePA” R package to perform enrichment analysis on the prioritized genes within Reactome pathways.^32^ Following the default settings, we considered the pathways as significant if *p*-values < 0.05 after being adjusted by the Benjamini-Hochberg method.

#### Observational analysis at population level

We used Cox proportional hazard model to calculate hazard ratios and 95% confidence intervals of incident RA associated with BRD2 genetype (rs206783). Model adjusted for age, sex, assessment centers, genotyping arrays and the first 10 genetic PC. The time scale of the cox model is based on the follow-up time, this was defined from the date of initial recruitment to the incident RA, date of death, loss of follow-up, or end of follow-up (31 0ctober 2022), whichever came first.

## Supplementary Information

**Document S1. Figures S1-S8**

**Figure S1.** Density histogram of p values in the rare variants burden tests from UKB exome sequencing studies

**Figure S2.** Sensitive analyses on the identified 18 proteins using Wald ratio or inverse-variance weighted two sample MR methods

**Figure S3.** Networks of three identified target genes (PP_H4_>0.8): *H1FX* (*H1-10*), *HAPLN4* and *W1SP1* (*CCN4*) in the colocalization analysis

**Figure S4.** Protein-protein interactions between current rheumatoid arthritis medications targets (JAK1, JAK2, JAK3, TNF, IL6R, CD20[MS4A1] and CD86) and identified potential drug targets

**Figure S5.** KEGG enrichment analysis on the candidate target genes that identified by three genetic informative approaches

**Figure S6.** Gene expression heatmap for candidate target genes based on GTEx V8 54 tissue types

**Figure S7.** BRD2 differential expression between rheumatoid arthritis patients (n=315) and healthy control (n=315) in CD14dim.CD16+ cell

**Figure S8.** Manhattan plot for PheWAS of BRD2 at gene level from AstraZeneca PheWAS Portal

**Document S2. Table S1-S17**

**Table S1**, Genes prioritized in MAGMA

**Table S2**, Genes prioritized in Pi

**Table S3**, Genes prioritized in PoPS

**Table S4**, Genes prioritized in DEPICT

**Table S5**, Genes prioritized in TWAS

**Table S6**, Gene intersections ascertained by the five gene priorization methods, Related to Figure 2A

**Table S7**, Enrichment of prioritized drug-target genes in the ATC codes, Related to Figure 2B

**Table S8**, Overlaps of these genes nominated by the omnibus gene prioritization ( ≥ 4 gene priorization methods jointly idetified) with drug target genes, Related to Figure 2C

**Table S9**, Top 5% significant genes associated with RA from rare variant burden test results computed on WES data in UKB

**Table S10**, Enrichment of prioritized drug-target genes in the ATC codes, based on prioritized genes from Table S9

**Table S11**, Associations of 1921 plasma proteins with the risk of rheumatoid arthritis using SMR & HEIDI methods, Related to Figure 3

**Table S12**. Sensitive analyses on the identified 18 proteins using Wald ratio or inverse-variance weighted two sample MR methods, Related to Figure S1

**Table S13**, Drug candidates for drug-target proteins with MR evidence, Related to Figure 4

**Table S14**, Overlaps of the prioritized genes or causal proteins with genes from the druggable genome

**Table S15,** Pathways significantly enriched in the genes prioritized by three approaches, related to Figure 5

**Table S16**, Phenotypes associated with BRD2 at variant level by PheWAS analysis, related to Figure 6

**Table S17**, Phenotypes associated with rs206783 (cis-pQTL for BRD2) in FinnGen

## References

1. Smolen JS, Aletaha D, McInnes IB. Rheumatoid arthritis. Lancet. 2016;388(10055):2023–2038. doi:10.1016/S0140-6736(16)30173-8

2. Finckh A, Gilbert B, Hodkinson B, et al. Global epidemiology of rheumatoid arthritis. Nat Rev Rheumatol. 2022;18(10):591–602. doi:10.1038/s41584-022-00827-y

3. Okada Y, Wu D, Trynka G, et al. Genetics of rheumatoid arthritis contributes to biology and drug discovery. Nature. 2014;506(7488):376–381. doi:10.1038/nature12873

4. GBD 2021 Rheumatoid Arthritis Collaborators. Global, regional, and national burden of rheumatoid arthritis, 1990-2020, and projections to 2050: a systematic analysis of the Global Burden of Disease Study 2021. Lancet Rheumatol. 2023;5(10):e594–e610. doi:10.1016/S2665-9913(23)00211-4

5. Alivernini S, Firestein GS, McInnes IB. The pathogenesis of rheumatoid arthritis. Immunity. 2022;55(12):2255–2270. doi:10.1016/j.immuni.2022.11.009

6. Smolen JS, Landewe RBM, Bijlsma JWJ, et al. EULAR recommendations for the management of rheumatoid arthritis with synthetic and biological disease-modifying antirheumatic drugs: 2019 update. Ann Rheum Dis. 2020;79(6):685–99. doi: 10.1136/annrheumdis-2019-216655.

7. Hsieh PH, Wu O, Geue C, McIntosh E, McInnes IB, Siebert S. Economic burden of rheumatoid arthritis: a systematic review of literature in biologic era. Ann Rheum Dis. 2020;79(6):771–777. doi:10.1136/annrheumdis-2019-216243

8. Putrik P, Ramiro S, Kvien TK, et al. Inequities in access to biologic and synthetic DMARDs across 46 european countries. Ann Rheum Dis. 2014;73(1):198–206. doi: 10.1136/annrheumdis-2012-202603.

9. Nelson MR, Tipney H, Painter JL, et al. The support of human genetic evidence for approved drug indications. Nat Genet. 2015;47(8):856–860. doi:10.1038/ng.3314

10. Trajanoska K, Bhérer C, Taliun D, Zhou S, Richards JB, Mooser V. From target discovery to clinical drug development with human genetics. Nature. 2023;620(7975):737–745. doi:10.1038/s41586-023-06388-8

11. Eric Vallabh Minikel, Jeffery L Painter, Coco Chengliang Dong, Matthew R. Nelson. Refining the impact of genetic evidence on clinical success. medRxiv 2023.06.23.23291765; doi: 10.1101/2023.06.23.23291765

12. Sekine C, et al. Successful treatment of animal models of rheumatoid arthritis with small-molecule cyclin-dependent kinase inhibitors. J Immunol. 2008;180:1954–1961.

13. Reay WR, Cairns MJ. Advancing the use of genome-wide association studies for drug repurposing. Nat Rev Genet. 2021;22(10):658–671. doi:10.1038/s41576-021-00387-z

14. Võsa U, Claringbould A, Westra HJ, et al. Large-scale cis- and trans-eQTL analyses identify thousands of genetic loci and polygenic scores that regulate blood gene expression. Nat Genet. 2021;53(9):1300–1310. doi:10.1038/s41588-021-00913-z

15. Fang H; ULTRA-DD Consortium, De Wolf H, et al. A genetics-led approach defines the drug target landscape of 30 immune-related traits. Nat Genet. 2019;51(7):1082–1091. doi:10.1038/s41588-019-0456-1

16. Zhao JH, Stacey D, Eriksson N, et al. Genetics of circulating inflammatory proteins identifies drivers of immune-mediated disease risk and therapeutic targets. Nat Immunol. 2023;24(9):1540–1551. doi:10.1038/s41590-023-01588-w

17. Ferkingstad E, Sulem P, Atlason BA, et al. Large-scale integration of the plasma proteome with genetics and disease. Nat Genet. 2021;53(12):1712–1721. doi:10.1038/s41588-021-00978-w

18. Folkersen L, Gustafsson S, Wang Q, et al. Genomic and drug target evaluation of 90 cardiovascular proteins in 30,931 individuals. Nat Metab. 2020;2(10):1135–1148. doi:10.1038/s42255-020-00287-2

19. Backman JD, Li AH, Marcketta A, et al. Exome sequencing and analysis of 454,787 UK Biobank participants. Nature. 2021;599(7886):628–634. doi:10.1038/s41586-021-04103-z

20. Szustakowski JD, Balasubramanian S, Kvikstad E, et al. Advancing human genetics research and drug discovery through exome sequencing of the UK Biobank. Nat Genet. 2021;53(7):942–948. doi:10.1038/s41588-021-00885-0

21. Sadler MC, Auwerx C, Deelen P, Kutalik Z. Multi-layered genetic approaches to identify approved drug targets. Cell Genom. 2023;3(7):100341. doi:10.1016/j.xgen.2023.100341

22. Namba S, Konuma T, Wu KH, Zhou W; Global Biobank Meta-analysis Initiative, Okada Y. A practical guideline of genomics-driven drug discovery in the era of global biobank meta-analysis. Cell Genom. 2022;2(10):100190. doi:10.1016/j.xgen.2022.100190

23. Ishigaki K, Sakaue S, Terao C, et al. Multi-ancestry genome-wide association analyses identify novel genetic mechanisms in rheumatoid arthritis. Nat Genet. 2022;54(11):1640–1651.

24. de Leeuw CA, Mooij JM, Heskes T, Posthuma D. MAGMA: generalized gene-set analysis of GWAS data. PLoS Comput Biol. 2015;11(4):e1004219. doi:10.1371/journal.pcbi.1004219

25. Pers TH, Karjalainen JM, Chan Y, et al. Biological interpretation of genome-wide association studies using predicted gene functions. Nat Commun. 2015;6:5890. doi:10.1038/ncomms6890

26. Weeks EM, Ulirsch JC, Cheng NY, et al. Leveraging polygenic enrichments of gene features to predict genes underlying complex traits and diseases. Nat Genet. 2023;55(8):1267–1276. doi:10.1038/s41588-023-01443-6

27. Mancuso N, Freund MK, Johnson R, et al. Probabilistic fine-mapping of transcriptome-wide association studies. Nat Genet. 2019;51(4):675–682. doi:10.1038/s41588-019-0367-1

28. Sakaue S, Okada Y. GREP: genome for REPositioning drugs. Bioinformatics. 2019;35(19):3821–3823. doi:10.1093/bioinformatics/btz166

29. Zheng J, Haberland V, Baird D, et al. Phenome-wide Mendelian randomization mapping the influence of the plasma proteome on complex diseases. Nat Genet. 2020;52(10):1122–1131. doi:10.1038/s41588-020-0682-6

30. Giambartolomei C, Vukcevic D, Schadt EE, et al. Bayesian test for colocalisation between pairs of genetic association studies using summary statistics. PLoS Genet. 2014;10(5):e1004383. doi:10.1371/journal.pgen.1004383

31. Szklarczyk D, Gable AL, Lyon D, et al. STRING v11: protein-protein association networks with increased coverage, supporting functional discovery in genome-wide experimental datasets. Nucleic Acids Res. 2019;47(D1):D607–D613. doi:10.1093/nar/gky1131

32. Yu G, He QY. ReactomePA: an R/Bioconductor package for reactome pathway analysis and visualization. Mol Biosyst. 2016;12(2):477–479. doi:10.1039/c5mb00663e

33. Ding Q, Hu W, Wang R, et al. Signaling pathways in rheumatoid arthritis: implications for targeted therapy. Signal Transduct Target Ther. 2023;8(1):68. doi:10.1038/s41392-023-01331-9

34. Zhang F, Wei K, Slowikowski K, et al. Defining inflammatory cell states in rheumatoid arthritis joint synovial tissues by integrating single-cell transcriptomics and mass cytometry. Nat Immunol. 2019;20(7):928–942. doi:10.1038/s41590-019-0378-1

35. Bycroft C, Freeman C, Petkova D, et al. The UK Biobank resource with deep phenotyping and genomic data. Nature. 2018;562(7726):203–209. doi:10.1038/s41586-018-0579-z

36. Fraenkel L, Bathon JM, England BR, et al. 2021 American College of Rheumatology Guideline for the Treatment of Rheumatoid Arthritis. Arthritis Rheumatol. 2021;73(7):1108–1123. doi:10.1002/art.41752

37. Eyre S, Bowes J, Diogo D, et al. High-density genetic mapping identifies new susceptibility loci for rheumatoid arthritis. Nat Genet. 2012;44(12):1336–1340. doi:10.1038/ng.2462

38. Zhao H, Rasheed H, Nøst TH, et al. Proteome-wide Mendelian randomization in global biobank meta-analysis reveals multi-ancestry drug targets for common diseases. Cell Genom. 2022;2(11):100195. doi:10.1016/j.xgen.2022.100195

39. Folkersen L, Gustafsson S, Wang Q, et al. Genomic and drug target evaluation of 90 cardiovascular proteins in 30,931 individuals. Nat Metab. 2020;2(10):1135–1148. doi:10.1038/s42255-020-00287-2

40. Sun BB, Maranville JC, Peters JE, et al. Genomic atlas of the human plasma proteome. Nature. 2018;558(7708):73–79. doi:10.1038/s41586-018-0175-2

41. Wu Y, Zeng J, Zhang F, Zhu Z, Qi T, Zheng Z, et al. Integrative analysis of omics summary data reveals putative mechanisms underlying complex traits. Nat Commun. 2018;9(1):918. doi: 10.1038/s41467-018-03371-0.

42. Chen Y, Sun W, Gao R, et al. The role of high mobility group box chromosomal protein 1 in rheumatoid arthritis. Rheumatology (Oxford). 2013;52(10):1739–1747. doi:10.1093/rheumatology/ket134

43. Wishart DS, Feunang YD, Guo AC, et al. DrugBank 5.0: a major update to the DrugBank database for 2018. Nucleic Acids Res. 2018;46(D1):D1074–D1082. doi:10.1093/nar/gkx1037

44. Chen X, Ji ZL, Chen YZ. TTD: Therapeutic Target Database. Nucleic Acids Res. 2002;30(1):412–415. doi:10.1093/nar/30.1.412

45. Whirl-Carrillo M, Huddart R, Gong L, et al. An Evidence-Based Framework for Evaluating Pharmacogenomics Knowledge for Personalized Medicine. Clin Pharmacol Ther. 2021;110(3):563–572. doi:10.1002/cpt.2350

46. Ochoa D, Hercules A, Carmona M, et al. Open Targets Platform: supporting systematic drug-target identification and prioritisation. Nucleic Acids Res. 2021;49(D1):D1302–D1310. doi:10.1093/nar/gkaa1027

47. Klein K, Kabala PA, Grabiec AM, et al. The bromodomain protein inhibitor I-BET151 suppresses expression of inflammatory genes and matrix degrading enzymes in rheumatoid arthritis synovial fibroblasts. Ann Rheum Dis. 2016;75(2):422–429. doi:10.1136/annrheumdis-2014-205809

48. Tsou PS, Sawalha AH. Glycoprotein nonmetastatic melanoma protein B: A key mediator and an emerging therapeutic target in autoimmune diseases. FASEB J. 2020;34(7):8810–8823. doi:10.1096/fj.202000651

49. Finan C, Gaulton A, Kruger FA, et al. The druggable genome and support for target identification and validation in drug development. Sci Transl Med. 2017;9(383):eaag1166. doi:10.1126/scitranslmed.aag1166

50. Hu, X., li, J., Fu, M., et al. The JAK/STAT signaling pathway: from bench to clinic. Sig Transduct Target Ther 6, 402 (2021). doi:10.1038/s41392-021-00791-1

51. Wu C, Tan S, Liu L, et al. Transcriptome-wide association study identifies susceptibility genes for rheumatoid arthritis. Arthritis Res Ther. 2021;23(1):38. doi:10.1186/s13075-021-02419-9

52. Maaten, Lvander & Hinton, G. Visualizing Data using t-SNE. J. Mach. Learn. Res. 9, 2579–2605 (2008).

53. Chen H, Xu J, Wei S, et al. RABC: Rheumatoid Arthritis Bioinformatics Center. Nucleic Acids Res. 2023;51(D1):D1381–D1387. doi:10.1093/nar/gkac850

54. Wang Q, Dhindsa RS, Carss K, Harper AR, Nag A, Tachmazidou I, Vitsios D, Deevi SVV, Mackay A, Muthas D, et al. Rare variant contribution to human disease in 281,104 UK Biobank exomes. Nature. 2021;597(7877):527–532.

55. Klein K. Bromodomain protein inhibition: a novel therapeutic strategy in rheumatic diseases. RMD Open. 2018;4:e000744. doi: 10.1136/rmdopen-2018-000744

56. Tyrsin D, Chuvpilo S, Matskevich A, et al. From TGN1412 to TAB08: the return of CD28 superagonist therapy to clinical development for the treatment of rheumatoid arthritis. Clin Exp Rheumatol. 2016;34(4 Suppl 98):45–48.

57. Toh ML, Bonnefoy JY, Accart N, et al. Bone- and cartilage-protective effects of a monoclonal antibody against colony-stimulating factor 1 receptor in experimental arthritis. Arthritis Rheumatol. 2014;66(11):2989–3000. doi:10.1002/art.3862

58. Genovese MC, Hsia E, Belkowski SM, et al. Results from a Phase IIA Parallel Group Study of JNJ-40346527, an Oral CSF-1R Inhibitor, in Patients with Active Rheumatoid Arthritis despite Disease-modifying Antirheumatic Drug Therapy. J Rheumatol. 2015;42(10):1752–1760. doi:10.3899/jrheum.141580

59. Schmidt AF, Finan C, Gordillo-Marañón M, et al. Genetic drug target validation using Mendelian randomisation. Nat Commun. 2020;11(1):3255. doi:10.1038/s41467-020-16969-0

60. Cao Y, Yang Y, Hu Q, Wei G. Identification of potential drug targets for rheumatoid arthritis from genetic insights: a Mendelian randomization study. J Transl Med. 2023;21(1):616. doi:10.1186/s12967-023-04474-z

61. Wang F, Chen FF, Gao WB, et al. Identification of citrullinated peptides in the synovial fluid of patients with rheumatoid arthritis using LC-MALDI-TOF/TOF. Clin Rheumatol. 2016;35(9):2185–2194. doi:10.1007/s10067-016-3247-4

62. Baron, R., Kneissel, M. WNT signaling in bone homeostasis and disease: from human mutations to treatments. Nat Med 19, 179–192 (2013). 10.1038/nm.3074

63. Li X, Shen A, Zhao Y, Xia J. Mendelian Randomization Using the Druggable Genome Reveals Genetically Supported Drug Targets for Psychiatric Disorders. Schizophr Bull. 2023;49(5):1305–1315. doi:10.1093/schbul/sbad100

64. Wang Q, Yang C, Gelernter J, Zhao H. Pervasive pleiotropy between psychiatric disorders and immune disorders revealed by integrative analysis of multiple GWAS. Hum Genet. 2015;134(11-12):1195–1209. doi:10.1007/s00439-015-1596-8

65. Eischer N, Arnold M, Mayer A. Emerging roles of BET proteins in transcription and co-transcriptional RNA processing. Wiley Interdiscip Rev RNA. 2023;14(1):e1734. doi:10.1002/wrna.1734

66. Gilan O, Rioja I, Knezevic K, et al. Selective targeting of BD1 and BD2 of the BET proteins in cancer and immunoinflammation. Science. 2020;368(6489):387–394. doi:10.1126/science.aaz8455

67. Zhu, Z., Zhang, F., Hu, H. et al. Integration of summary data from GWAS and eQTL studies predicts complex trait gene targets. Nat Genet 48, 481–487 (2016). 10.1038/ng.3538

68. Zhang T, Klein A, Sang J, Choi J, Brown KM. ezQTL: A Web Platform for Interactive Visualization and Colocalization of QTLs and GWAS Loci. Genomics Proteomics Bioinformatics. 2022;20(3):541–548. doi:10.1016/j.gpb.2022.05.004

69. Warde-Farley D, Donaldson SL, Comes O, et al. The GeneMANIA prediction server: biological network integration for gene prioritization and predicting gene function. Nucleic Acids Res. 2010;38(Web Server issue):W214–W220. doi:10.1093/nar/gkq537

