## Supplemental Figure S1-S8 for "Identifying therapeutic targets for rheumatoid arthritis by genomics-driven integrative approaches"

J Zhang et al.

Corresponding to DQ Ye

**Table of contents**

**Supplementary Figures**

**Figure S1**. Density histogram of p values in the rare variants burden tests from UKB exome sequencing studies

**Figure S2**. Sensitive analyses on the identified 18 proteins using Wald ratio or inverse-variance weighted two sample MR methods

**Figure S3**. Networks of three identified target genes (PP_H4_>0.8): *H1FX* (*H1-10/H1X*), *HAPLN4* and *W1SP1* (*CCN4*) in the colocalization analysis

**Figure S4**. Protein-protein interactions between current rheumatoid arthritis medications targets (JAK1, JAK2, JAK3, TNF, IL6R, CD20[MS4A1] and CD86) and identified potential drug targets

**Figure S5**. KEGG enrichment analysis on the candidate target genes that identified by three genetic informative approaches

**Figure S6**. Gene expression heatmap for candidate target genes based on GTEx V8 54 tissue types

**Figure S7**. BRD2 differential expression between rheumatoid arthritis patients (n=315) and healthy control (n=315) in CD14dim.CD16+ cell

**Figure S8**. Manhattan plot for PheWAS of BRD2 at gene level from AstraZeneca PheWAS Portal

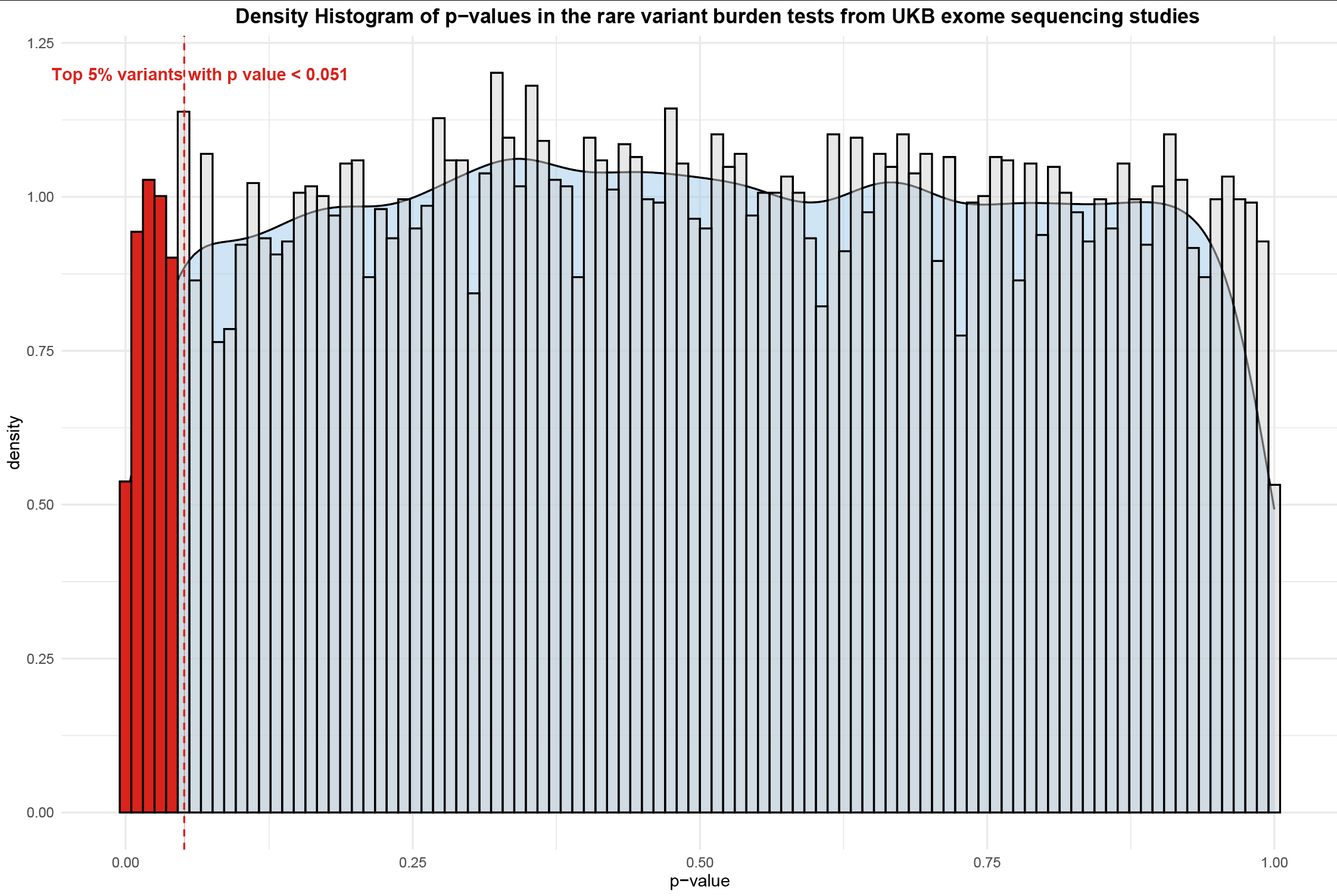

**Figure S1**. Density histogram of p values in the rare variants burden tests from UKB exome sequencing studies (GWAS catalog access ID: GCST90085490).

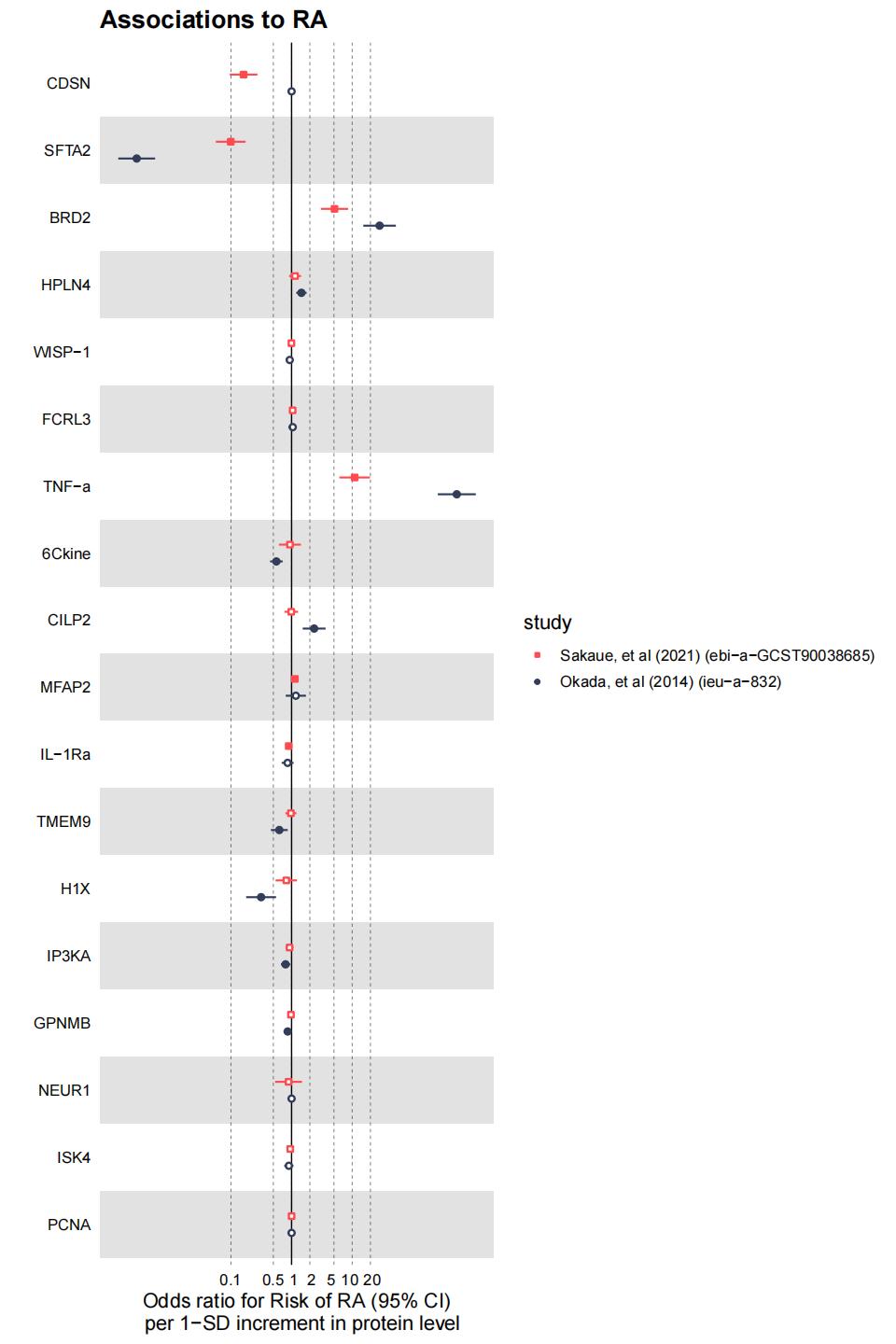

**Figure S2**. Sensitive analyses on the identified 18 proteins using Wald ratio or inverse-variance weighted two sample MR methods. A solid dot indicates that the p-value is less than 0.05, and otherwise, it is greater than 0.05.

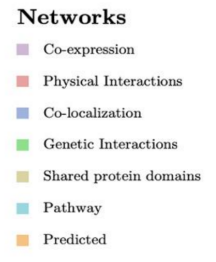

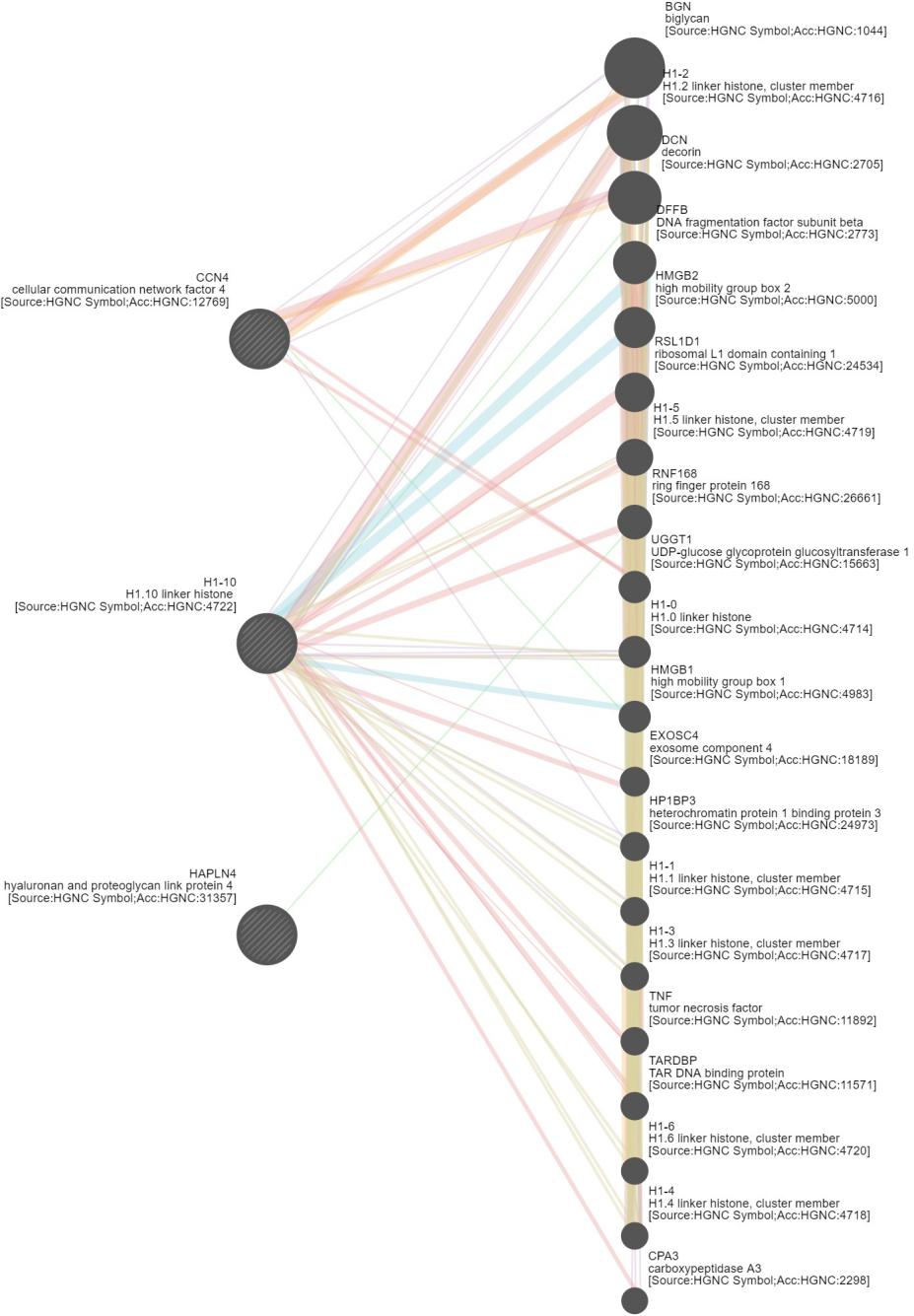

**Figure S3**. Networks of three identified target genes (PP_H4_>0.8): *H1FX* (H1-10), *HAPLN4* and *W1SP1* (CCN4) in the colocalization analysis. The network prediction was based on an online tool: GeneMANIA (http://www.genemania.org).

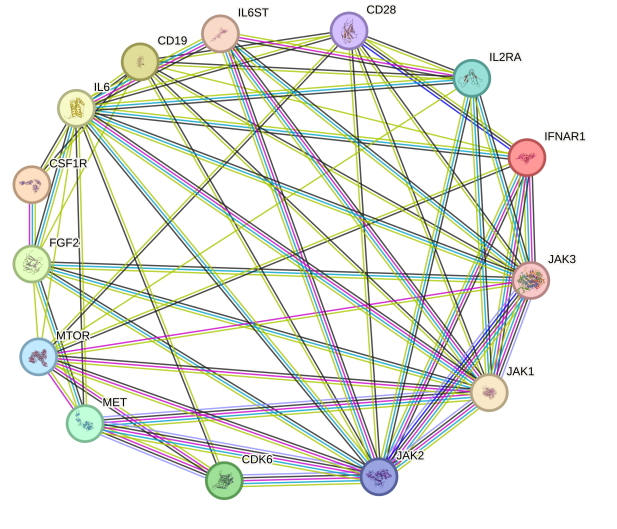

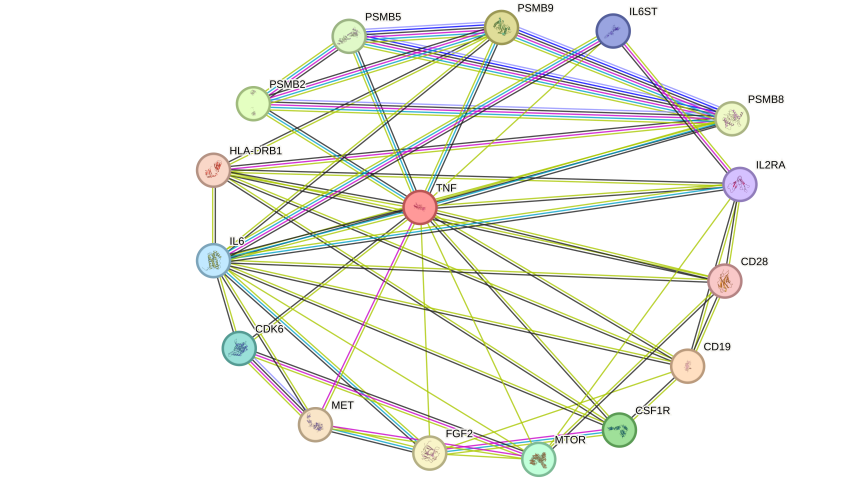

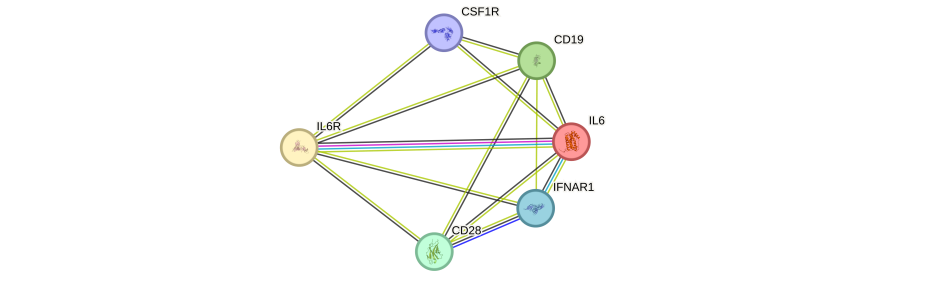

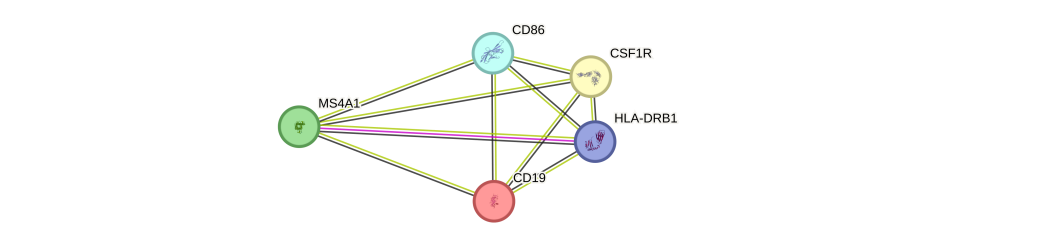

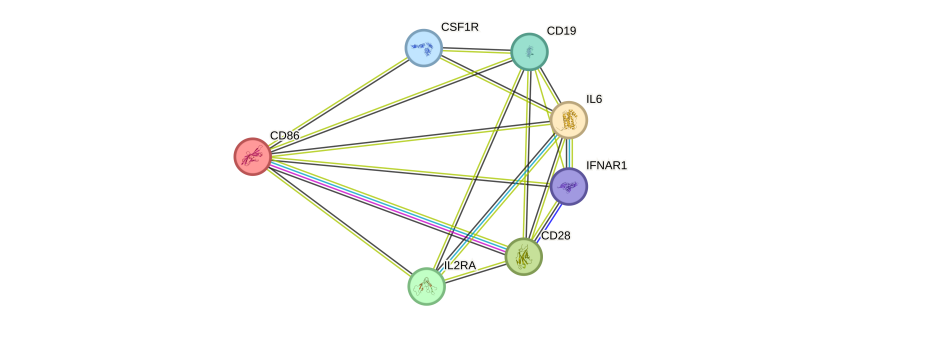

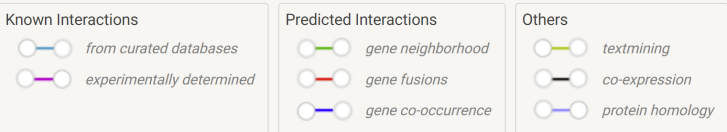

**Figure S4**. Protein-protein interactions between current rheumatoid arthritis medications targets (JAK1, JAK2, JAK3, TNF, IL6R, CD20[MS4A1] and CD86) and identified potential drug targets.

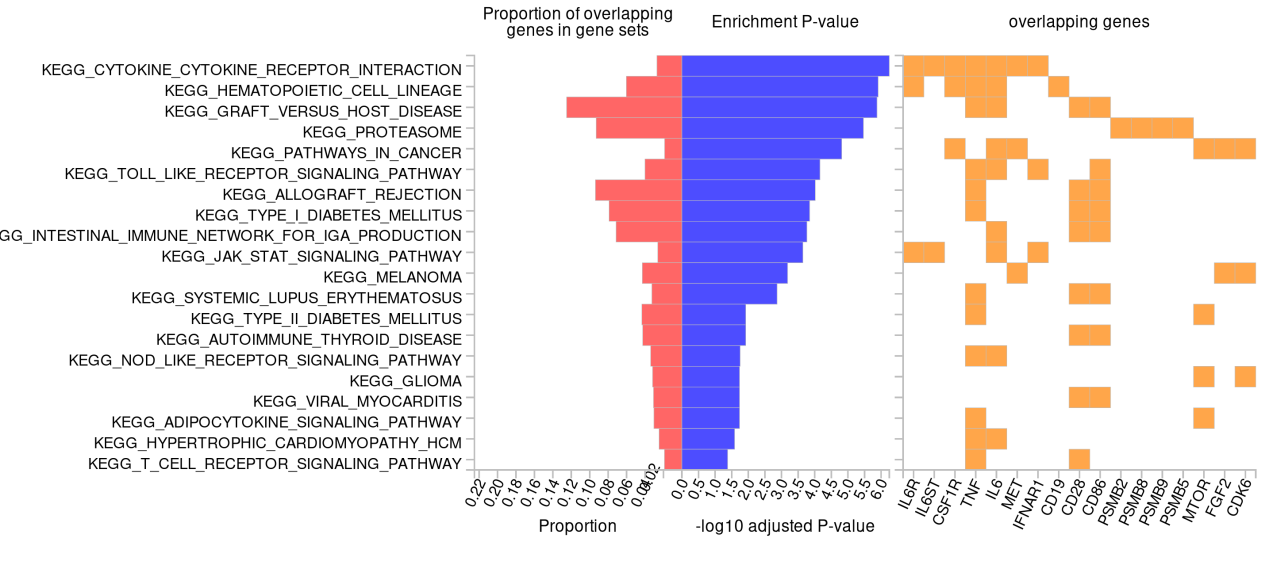

**Figure S5**. KEGG enrichment analysis ( GENE2FUNC, https://fuma.ctglab.nl/gene2func) on the candidate target genes that identified by three genetic informative approaches.

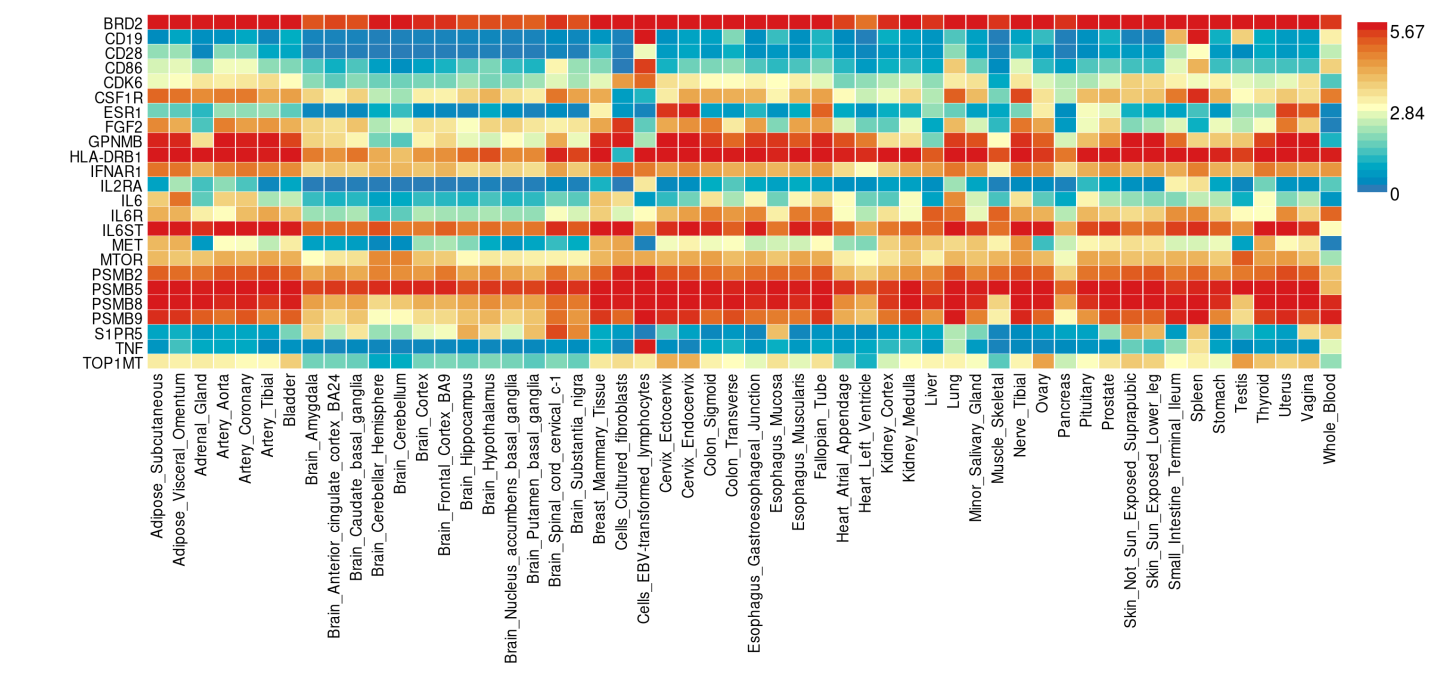

**Figure S6**. Gene expression heatmap for candidate target genes based on GTEx V8 54 tissue types.

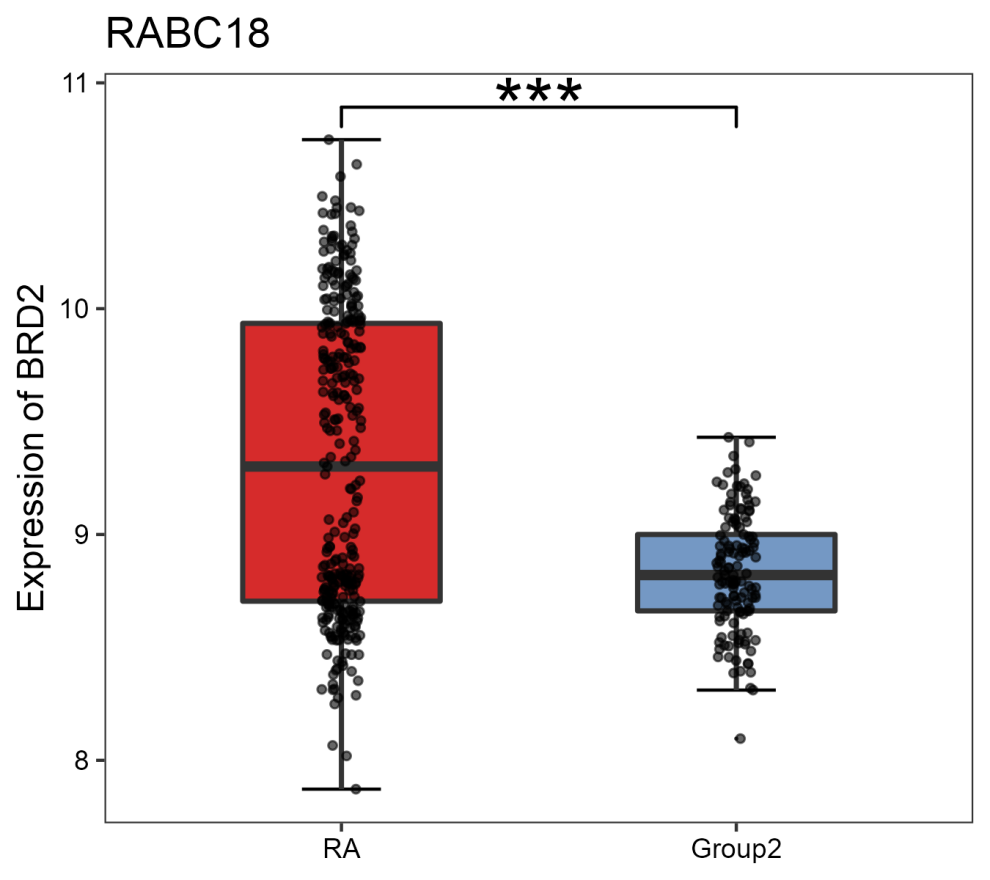

**Figure S7**. BRD2 differential expression between rheumatoid arthritis patients (n=315) and healthy control (n=315) in CD14dim.CD16+ cell. (the differential expression analysis results from http://www.onethird-lab.com/RABC/index.php/Index/dataset_details?id=RABC18)

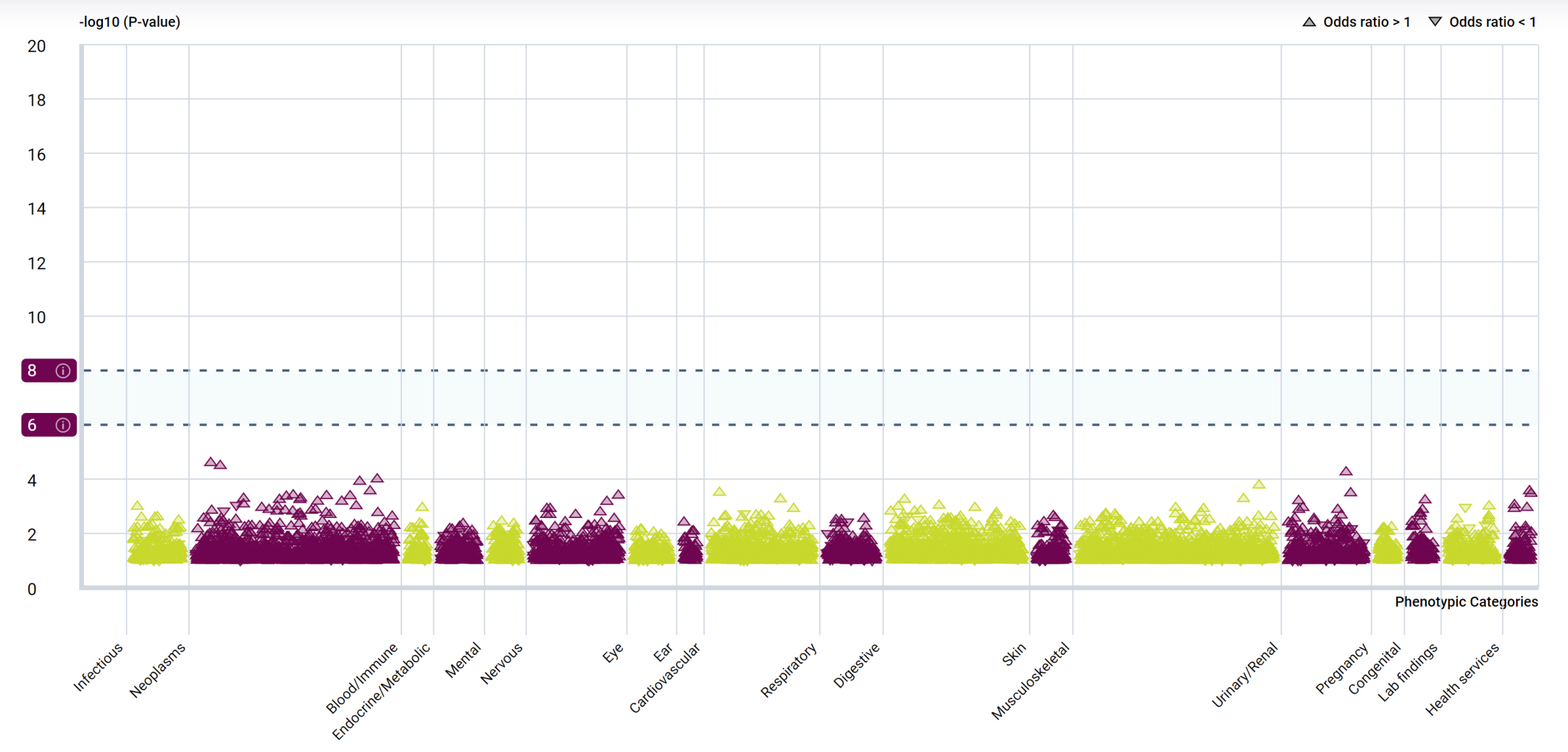

**Figure S8**. Manhattan plot for PheWAS of BRD2 at gene level from AstraZeneca PheWAS Portal (https://azphewas.com/).
